# CDK4/6 and SHP2 mediate BRAF/MEK inhibitor resistance in Class 2 and 3 BRAF mutant cancers

**DOI:** 10.1101/2024.07.28.24311101

**Authors:** April A. N. Rose, Jennifer Maxwell, Emmanuelle Rousselle, Melody Riaud, Islam E. Elkholi, Chantel Mukonoweshuro, Marco Biondini, Erica Cianfarano, Isabel Soria-Bretones, Chantal Tobin, Meghan McGuire, Ian King, Tong Zhang, Trevor J. Pugh, Zaid Saeed Kamil, Frances A. Shepherd, Natasha Leighl, Albiruni Abdul Razak, Aaron Hansen, Sam Saibil, Philippe Bedard, Peter M. Siegel, Lillian L. Siu, David W. Cescon, Anna Spreafico

## Abstract

Class 2 and 3 non-V600E *BRAF* mutations are oncogenic drivers in many cancer types. Currently, there are no established targeted therapies with proven efficacy for cancers with non-V600E *BRAF* mutations. We developed the investigator-initiated, Phase II BEAVER clinical trial (NCT03839342) to evaluate the efficacy of BRAF and MEK inhibitors in patients with non-V600E *BRAF* mutations. The best objective response rate was 14% (3/21). By analyzing genomic data from patient tumors, circulating tumor DNA (ctDNA), patient-derived xenograft (PDX) models generated from enrolled patients, and functional genomics of Class 2 & 3 *BRAF* mutant cell lines, we discovered MAPK-dependent and independent mechanisms of intrinsic and acquired resistance to BRAF/MEK inhibition. These included the acquisition of new mutations in *NRAS, MAP2K1, RAF1,* and *RB* in ctDNA at the time of disease progression. We observed an enrichment for alterations in genes that regulate cell cycle progression amongst non-responders and increased expression of genes mediating cell cycle progression in tumors Class 2 *BRAF* mutant cell lines with acquired resistance to BRAF/MEK inhibitors. In Class 3 BRAF mutant cancers specifically, *PTPN11* (SHP2) was an essential gene. CDK4/6 and SHP2 were found to mediate intrinsic resistance to BRAF/MEK inhibition in Class 2 & 3 BRAF mutant tumors. Therapeutic strategies combining CDK4/6 or SHP2 inhibitors with BRAF/MEK inhibitors were more effective than BRAF/MEK inhibitors alone *in vitro* and *in vivo*, highlighting the need to explore therapeutic targets outside of the MAPK pathway in these hard-to-treat Class 2 & 3 *BRAF* mutant cancers.

## Introduction

*BRAF* is one of the most frequently mutated actionable oncogenes in cancer (1,2). Oncogenic Class 1 (V600) mutations in BRAF render this kinase constitutively active, leading to excessive Mitogen Activated Protein Kinase (MAPK) pathway activity, increased cell proliferation, and tumorigenesis. Combined inhibition of BRAF and its downstream kinase, MEK, is an effective therapeutic strategy for most cancer types with *BRAF* V600E mutations (3). However, non-V600 BRAF mutants account for approximately 30% of all oncogenic *BRAF* mutations in adult solid tumors (4), and currently there are no established targeted therapies for non-V600 BRAF mutant tumors.

There are a wide array of oncogenic non-V600 *BRAF* mutations that can be subdivided into Class 2 and 3 mutations (1). Unlike Class 1 BRAF mutants - which signal as monomers - Class 2 and 3 non-V600 BRAF mutants signal as dimers. Class 2 BRAF mutants form RAS-independent dimers with intermediate-to-high kinase activity (5), whereas Class 3 *BRAF* mutations confer low kinase activity but form RAS-dependent dimers with other RAF proteins to hyperactivate the downstream MEK and ERK kinases (6). Indeed, *RAS* mutations frequently co-occur with Class 3 *BRAF* mutations in cancer (4,6). In RAS wild-type tumors, the protein tyrosine phosphatase, SHP2, has been implicated as a key regulator of RAS activity that potentiates RAF dimerization (7,8). Once phosphorylated, ERK regulates cell cycle progression by inducing the transcription of Cyclin D1, promoting its assembly with the cyclin-dependent kinases, CDK4 and CDK6 (9). Cyclin-D-bound CDK4/6 and Cyclin-E-bound CDK2 kinases phosphorylate and inactivate the tumor suppressor RB (10). This step is essential for cells to progress through the G1-S phase of the cell cycle. Excessive cell-cycle progression is prevented in part by the tumor suppressor, p21, a p53-regulated cyclin-dependent kinase inhibitor protein, which inhibits the activity of CDK2 and in some contexts CDK4/6 (11). Therefore, in cells with loss-of-function *TP53* mutations, progression through the cell cycle is a less tightly controlled process. Of note, *TP53* mutations are also more likely to co-occur in tumors with Class 2 and 3 vs. Class 1 *BRAF* mutations (4).

*RAS* mutations can potentiate BRAF inhibitor-induced paradoxical activation of the MAPK pathway in BRAF wild-type tumors (12); thus, earlier studies proposed MEK inhibitor monotherapy for tumors with Class 2 and 3 *BRAF* mutations (13). However, one prospective clinical trial demonstrated a lack of clinical efficacy for trametinib monotherapy in these tumors (14). Subsequently, it was reported that BRAF monomer inhibitors do not promote paradoxical activation of the MAPK pathway even in RAS co-mutated non-V600E BRAF mutant tumors (15). Moreover, combined BRAF and MEK inhibition was more effective than MEK inhibitor monotherapy at inhibiting tumor growth in preclinical models of cancers with Class 2 or 3 *BRAF* mutations (15,16). A recent systematic review of primarily retrospective data revealed response rates as high as 50% in patients with Class 2 or 3 BRAF mutant solid tumors that were treated with combined BRAF + MEK inhibitors (17). We designed a Phase II clinical trial to prospectively evaluate the efficacy of <u>B</u>inimetinib (MEK inhibitor) and <u>E</u>ncorafenib (BRAF inhibitor) for the treatment of <u>A</u>dvanced solid tumors with non-<u>V</u>600<u>E</u> *B<u>R</u>AF* mutations (BEAVER trial). We also sought to characterize molecular mechanisms associated with treatment response and resistance in these tumors.

## Results

### Clinical characteristics and outcomes of patients enrolled on the BEAVER trial

The BEAVER trial was a Princess Margaret Cancer Centre investigator-initiated, Phase II clinical trial evaluating binimetinib and encorafenib (B+E) for the treatment of advanced solid tumors with non-V600E *BRAF* mutations, with a primary objective of overall response rate (ORR) by RECIST v1.1. Exploratory objectives included genomic and transcriptomic profiling to interrogate mechanisms of treatment response and resistance and development of patient-derived xenograft (PDX) models of non-V600E BRAF mutant cancers (**Figure 1a**). Twenty-seven patients were screened and 23 out of a planned 26 patients were enrolled in the BEAVER trial. The trial closed early due to poor accrual during the COVID-19 pandemic. Enrolled patients’ characteristics are described in **(Table S1)**. The tumor types included melanoma, colorectal and pancreaticobiliary (n=6 each), lung (n=2), and breast, uterine, and small bowel cancers (n=1 each). The median age was 59 years. Patients’ tumors had Class 1 (n=1), Class 2 (n=9), and Class 3 (n=13) *BRAF* mutations.

**Figure 1:**
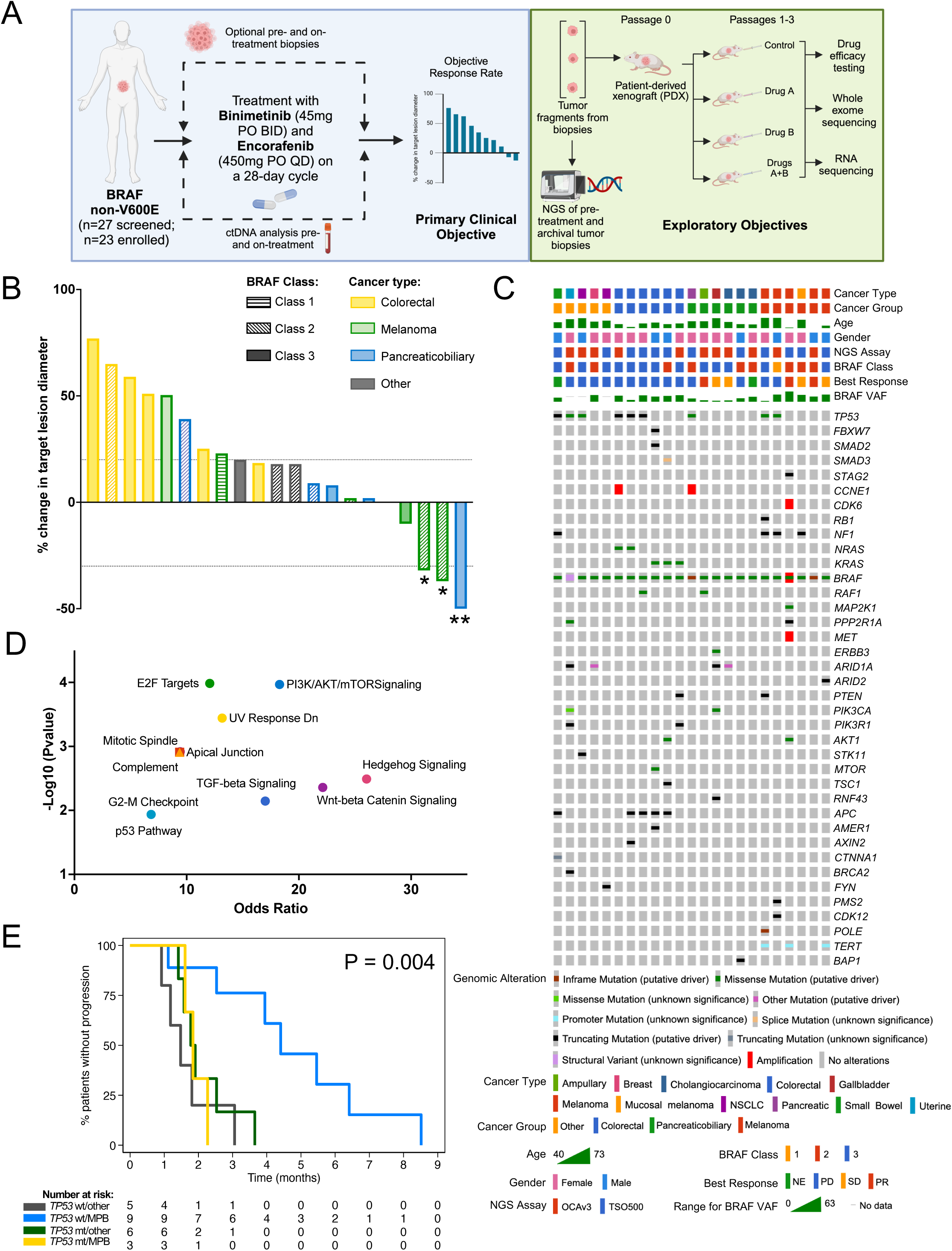
Clinical efficacy of binimetinib and encorafenib in patients with non-V600E *BRAF* mutations. **A)** Schematic of BEAVER Trial Study Design. **B)** Waterfall plot of tumor measurements assessed by RECIST 1.1. *unconfirmed PR; **confirmed PR. **C)** Oncoprint indicating all oncogenic or likely oncogenic alterations that were identified using either a OCAv3 or TSO500 NGS assay on archival or fresh tumor biopsies taken from all enrolled patients prior to initiating treatment on the study. **D)** The list of genes shown in the oncoprint was analyzed by Hallmark MSigDB pathways analysis to identify altered pathways that were enriched in this cohort. All enriched pathways that were Padj<0.05 are shown. **E)** Kaplan-Meier curve indicating the progression free survival of patients with either melanoma or pancreaticobiliary cancers vs. other cancer types that were either TP53 mutant or wildtype.

Twenty-one patients were evaluable for ORR. The best ORR was 14% (3/21), confirmed ORR was 5% (1/21) (**Figure 1b**). One patient with a Class 3 (*BRAF* D594G) ampullary cancer had a confirmed partial response (PR) and two melanoma patients with Class 2 *BRAF* mutations (K601E, G469S) experienced unconfirmed PRs. Four patients had stable disease (SD) as best response, two patients were non-evaluable (NE) for response, and 14 patients had progressive disease (PD) as best response. In the entire cohort, the median PFS was 2.3 months and the median OS was 6.1 months (**Figure S1**). This study did not meet the pre-specified criteria of 4/26 responses required for further investigation of this regimen in this patient population. Twenty-three patients were evaluable for safety. The adverse event profile of B+E **(Table S2)** was similar to what has previously been reported for this regimen (18,19). In this cohort, 22% of patients experienced Grade 3 treatment related adverse events. There were no Grade 4 or 5 treatment related adverse events and no new safety signals were identified.

We performed secondary analyses to investigate clinical and genomic biomarkers of response and resistance. The ORR and PFS did not differ significantly according to *BRAF* mutation Class (**Figure S2**). However, cancer type was associated with differences in response rate and PFS (**Figure S3a**). Moreover, patients with melanoma experienced longer PFS (4.0 months) than patients with or pancreaticobiliary (2.6 months) or colorectal or other tumor types (1.8 months; P=0.021; **Figure S3b**).

### Genomic characteristics of patients’ tumors

Targeted panel sequencing of relevant cancer driver genes was performed on archival tumors or fresh pre-treatment tumor biopsies for all patients enrolled on the BEAVER trial. Co-occurring pathogenic variants were identified in 21/23 (91%) tumors (**Figure 1c**). Pathway analysis of the co-altered genes revealed enrichment for alterations in multiple potentially actionable pathways (**Figure 1d**). In this cohort 12/23 (52%) patients had a co-occurring MAPK pathway activating mutation in: *NF1* (n=4), *KRAS* (n=3), *NRAS* (n=2), *RAF1* (n=2) or *MAP2K1* (n=1) genes at baseline. The most frequently co-altered gene was *TP53*, which was mutated in 9/23 (39%) tumors. All evaluable patients with tumor *TP53* mutations had PD as best response (**Figure 1c**). Patients with melanoma or pancreaticobiliary tumors that were *TP53* wild-type were significantly more likely to have SD or PR as best response (**Figure S3d**) and experienced longer PFS (4.4 months) compared to patients who had other cancer types or patients with melanoma/pancreaticobiliary cancer with *TP53* mutations (1.2-1.6 months; P=0.004) (**Figure 1e**).

### Development of pre-clinical models of BRAF/MEK inhibitor resistant Class 2 & 3 BRAF mutant cancers

The response rate to B+E treatment in this population of patients with non-V600E BRAF mutations was substantially lower, and the PFS was shorter, than it is for patients with *BRAF* V600 mutations (18,19). Therefore, we sought to identify mechanisms of resistance to B+E in cancers with Class 2 and 3 *BRAF* mutations. To do this, we developed patient-derived xenograft (PDX) models from patients enrolled on the BEAVER trial. Fresh tumor biopsy samples from 13 patients were implanted into NSG mice. The details of patients, biopsies and BRAF mutations are described in **(Table S3).** Of these, PDXs were successfully established from 9/13 (69%) patient samples (**Figure 2a**). Successful engraftment of a PDX was associated with an increased % change in target lesion size by RECIST 1.1 in the corresponding patient (**Figure S4a**). We performed whole exome sequencing on PDXs and confirmed the presence of a non-V600E BRAF mutation in 8/9 established PDXs (**Figure S4b**). Amongst the BRAF mutant PDXs, there was a high concordance of oncogenic alterations between the patient tumor and the established PDX (**Figure S4b**). We evaluated the ability of B+E to promote tumor growth inhibition in the non-V600E BRAF mutant PDXs *in vivo* (**Figure S4c**). We observed a significant indirect correlation between the amount of B+E induced tumor growth inhibition in PDXs *in vivo* with the change in target lesion size from the corresponding patients. PDXs that showed more tumor growth inhibition with B+E *in vivo* were derived from patients that experienced less growth in target lesion size during B+E treatment (**Figure 2b, Figure S4c**). To expand our panel of preclinical models of B+E-resistant non-V600E BRAF cancers, we separately generated four independent Class 2 BRAF mutant cancer cell lines with acquired resistance to B+E (**Figure 2c**). These included two melanoma (HMV-II, FM95), one breast cancer (MDA-MB-231), and one prostate cancer (22RV1) cell line. Cells were grown continuously in the presence of B+E - in a 1:5 ratio of binimetinib:encorafenib, to be consistent with the clinical drug dosing ratio - for several months. B+E resistance was confirmed by clonogenic assay (**Figure 2d**) and by comparing the IC50 of all parental/resistant pairs (**Figure 2e, Figure S5a**). Together, these models allowed us to study the intrinsic and adaptive mechanisms of resistance to B+E in Class 2 and 3 BRAF mutant cancers.

**Figure 2:**
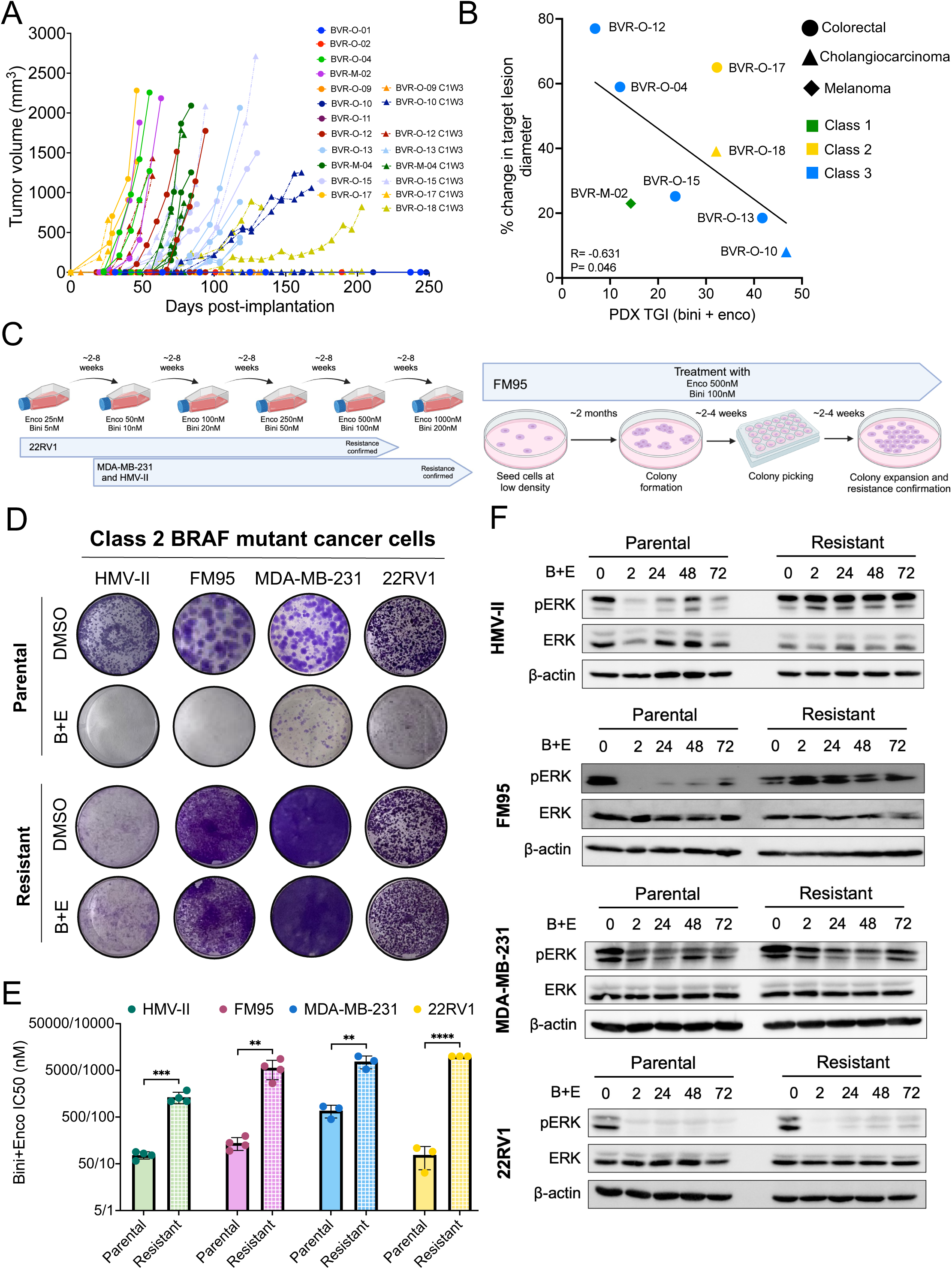
Generation and characterization of preclinical models of BRAF/MEK inhibitor resistant Class 2 and 3 BRAF mutant cancer. **A)** Tumor growth curves of P0 PDXs derived from patients enrolled on the BEAVER trial **B)** Correlation of tumor growth observed with Bini+Enco treatment in patients versus relative tumor growth inhibition achieved with bini+enco treatment in corresponding PDXs. Patient tumor growth measurement represents the % change in sum of target lesion diameter as per RECIST 1.1. R=-0.63; P=0.0467; Pearson correlation coefficient. **C)** Schematic of the two methods used to generate Bini+Enco resistant cell lines. Dose escalation protocol is shown on the left, and colony selection protocol is shown on the right. **D)** Representative crystal violet images from parental and resistant Class 2 BRAF cancer cell lines treated with DMSO or Bini+Enco (200nM Bini + 1000nM Enco) for 7-10 days. **E)** Bini+Enco IC50 values for each parental/resistant pair, N=3 biological replicates plotted. IC50s that were not achieved were represented as the maximal concentration tested (2000nM Bini + 10000nM Enco). Unpaired t-test *P<0.05, **P<0.01, ***P<0.001, ****P<0.0001. **F)** Representative immunoblots of MAPK pathway activity in the Class 2 BRAF mutant parental/resistant cancer cell lines. Cells were treated with Bini+Enco (100nM Bini + 500nM Enco) for 2, 24, 48, and 72hrs or DMSO (B+E 0) for 72 hours.

### Characterization of cell cycle alterations in BRAF/MEK inhibitor resistant Class 2 BRAF mutant cells

MAPK pathway reactivation is a common feature of BRAF/MEK inhibitor resistance in melanomas with Class 1 *BRAF* mutations (20,21). We sought to determine if this phenomenon is recapitulated in Class 2 BRAF mutant cells. Therefore, we assessed the relative degree of MAPK pathway activity (via ERK phosphorylation, pERK) in parental and resistant Class 2 cells. The Class 2 BRAF mutant resistant melanoma cells lines (HMV-II, FM95) displayed robust MAPK re-activation, despite treatment with B+E (**Figure 2f**). However, MAPK pathway inhibition was minimal in parental MDA-MB-231 breast cancer cells and remained unchanged in resistant cells. Conversely, B+E potently inhibited MAPK pathway activity in both parental and resistant 22RV1 prostate cancer cells. Our results demonstrate that Class 2 BRAF mutant cancers do not universally employ MAPK pathway reactivation as a resistance mechanism to BRAF/MEK inhibition. As the MAPK pathway regulates apoptosis, we evaluated the impact of B+E on apoptosis. B+E induced apoptosis in all Class 2 parental cell lines, and there was a significant reduction in B+E-induced apoptosis in the B+E-resistant melanoma cells. However, there was no significant difference in apoptosis levels between B+E-treated MDA-MB-231 and 22RV1 parental/resistant cells (**Figure S5b**).

The four pairs of parental/resistant cells were treated for 24hrs in the presence or absence of B+E and subjected to RNA sequencing (RNA-Seq) and analysis. In principal component analyses, the primary differential component was determined by cancer type rather than cell line resistance to BRAF/MEK inhibitors (**Figure 3a**). Gene Set Enrichment Analysis (GSEA) revealed 8 gene sets that were commonly enriched in at least 3/4 B+E-treated resistant cells compared to B+E-treated parental cell lines (**Figure 3b**, complete GSEA results in **Table S4**). This included two gene-sets (E2F targets & G2M Checkpoint) that were altered in the BEAVER trial patient tumors (**Figure 1d**) and responsible for mediating cell cycle progression. Additionally, B+E treatment potently suppressed expression of genes in the E2F Targets and G2M Checkpoint gene sets in Class 2 parental cells, but this effect was abrogated in resistant cells (**Figure 3c, 3d, Figure S6a**). Drug-induced MAPK pathway inhibition was confirmed by evaluating the MAPK pathway activity score (MPAS, (22), **Figure S6a,b**) and was observed in all Class 2 cell lines and did not differ by cancer type (**Figure S6c**). However, B+E did more potently suppress expression of the E2F/G2M gene sets in melanoma vs. non-melanoma cell lines (**Figure S6c**). Thus, we hypothesized that cell cycle progression may be altered as a resistance mechanism in our B+E-resistant cells.

**Figure 3:**
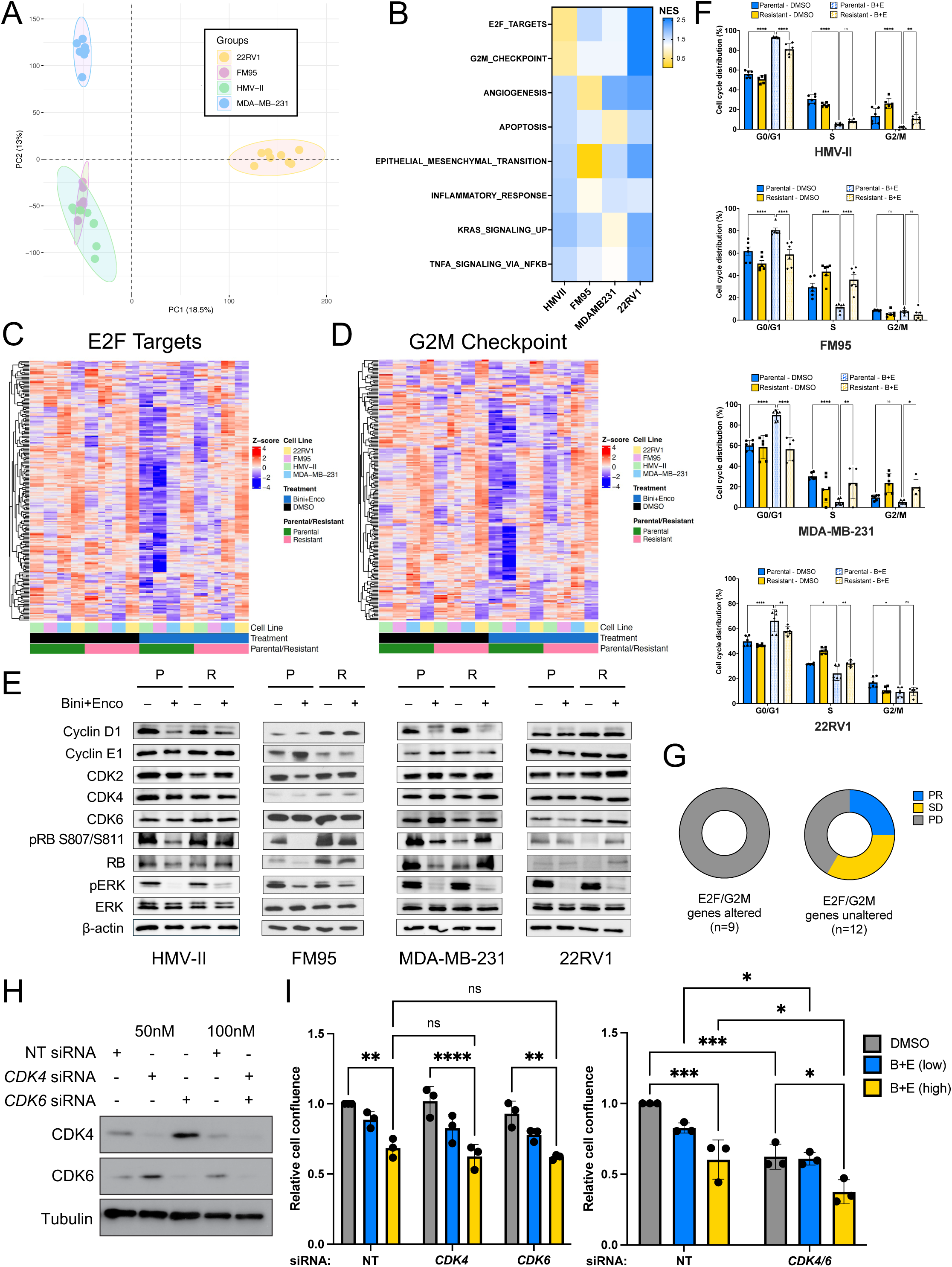
Identification of altered cell cycle regulation in resistant Class 2 BRAF mutant cells. **A)** Principal Component Analysis plot of the Bini+Enco and DMSO treated Parental/Resistant non-V600 BRAF Class 2 cell lines. **B)** Normalized Enrichment Score from GSEA comparing each Bini+Enco treated Parental/Resistant non-V600 BRAF Class 2 cell lines. Plotted Gene Sets from the top 15 most enriched common gene sets between the 4 cell lines. **C)** Heatmap of the Hallmark MSigDB E2F Targets gene set (genes = 198, baseMean>0) in the Bini+Enco and DMSO treated Parental/Resistant non-V600 BRAF Class 2 cell lines. **D)** Heatmap of the Hallmark MSigDB G2M Checkpoint gene set (genes = 196, baseMean>0) in the Bini+Enco and DMSO treated Parental/Resistant non-V600 BRAF Class 2 cell lines. **E)** Immunoblot of cell cycle proteins from synchronized cells treated with DMSO or Bini (100nM) + Enco (500nM) for FM95 and 22RV1 and Bini (200nM) + Enco (1000nM) for HMV-II and MDA-MB-231 for 48hrs. **F)** Bar graph of the cell cycle phase distribution based on propidium iodide flow cytometry cell cycle analysis. Cells were synchronized overnight and treated with DMSO or Bini (200nM) + Enco (1000nM) for 24hrs. Differences were evaluated with a 2-way ANOVA with Tukey correction for multiple comparisons. **G)** Responses to Bini+Enco treatment amongst BEAVER trial patients, stratified according to the presence or absence of oncogenic alterations in genes within the E2F Targets or G2M Checkpoint gene sets (P=0.0226). **H)** MDA-MB-231 cells were transfected with the indicated concentrations of siRNA and analyzed by immunoblotting at experimental endpoint. **I)** Proliferation assay comparing the response of siRNA-transfected MDA-MB-231 cells to either low (Bini 25nM + Enco 125nM) or high dose (Bini 200nM + Enco 1000nM) drug treatment. Two-way ANOVA, Tukey correction *P<0.05, **P<0.01, ***P<0.001, ****P<0.0001.

Cell cycle progression is tightly regulated by the tumor supressor RB. RB is phosphorylated by cyclin-dependent kinases, CDK2, CDK4 and CDK6, liberating E2F to promote transcription of E2F target genes required for G1-S phase transition. CDK activity is regulated by cyclin D (CDK4/6) and cyclin E (CDK2). Therefore we asked whether resistant cells were characterized by changes in these proteins. In synchronized cells, a 48hr B+E treatment potently suppressed cyclin D protein (observed in FM95 after 24hr treatment, data not shown) and phosphorylated RB (pRB) levels in parental Class 2 BRAF mutant melanoma cells, to a lesser extent in MDA-MB-231 cells but did not suppress cyclin D/pRB in 22RV1 cells (**Figure 3e**). In resistant cells, B+E treatment was not sufficient to inhibit RB phosphorylation. We did not observe any substantial or consistent differences in CDK protein expression level between parental and resistant cell lines (**Figure 3e**). Next, we assessed cell cycle distribution in parental and resistant cells. B+E treatment significantly increased G0/G1 arrest in all parental Class 2 cells and this B+E-dependent effect was abrogated in all resistant cells (**Figure 3f**). B+E reduced the percentage of cells in S and/or G2/M-phase in all parental cell lines, but this effect was diminished in resistant cells (**Figure 3f, Figure S6d**). Moreover, BEAVER trial patients with tumors harboring co-alterations in genes within the E2F Targets (*CCNE1*, *PMS2*, *BRCA2*, *POLE* and *TP53*) and/or G2M Checkpoints (*SMAD3, BRCA2*, and *POLE* and *TP53*) gene sets were intrinsically resistant to B+E treatment (**Figure 3g**). These data highlight the potential therapeutic value of targeting mediators of cell cycle progression to enhance the efficacy of B+E in Class 2 BRAF mutant cancers.

### Evaluating BRAF/MEK/CDK4/6 inhibitor combinations in Class 2 BRAF mutant cancers

CDK4/6 kinases are critical in regulating G1-S-phase transition and represent actionable therapeutic targets in cancer. Therefore, we tested whether CDK4 and/or CDK6 are required to mediate resistance to B+E in Class 2 BRAF mutant cancer cells. In MDA-MB-231 cells, CDK4 and CDK6 were knocked down by siRNA alone or in combination (**Figure 3h**). Knockdown of either CDK4 or CDK6 alone did not inhibit cellular proliferation or potentiate the growth-inhibition effect mediated by B+E. However, simultaneous knockdown of CDK4 and CDK6 inhibited the proliferation of DMSO-treated cells. Notably, the combined knockdown further potentiated cell growth inhibition mediated by B+E at low or high doses (**Figure 3i**). Together, these data led us to investigate the therapeutic benefit of a combined CDK4/6 inhibition with BRAF and MEK inhibition in Class 2 BRAF mutant cancers.

We assessed the efficacy of the CDK4/6 inhibitor, palbociclib, for inhibiting cell proliferation of Class 2 BRAF mutant parental cancer cells, when used alone or in combination with B+E. In line with our CDK4/6 knockdown experiments, treatment with palbociclib alone inhibited cell survival in 3/4 Class 2 BRAF mutant cell lines. The triple therapy combination (B+E+palbociclib; B+E+P) was more effective at inhibiting cell growth vs. B+E in all four Class 2 cell lines tested (MDA-MB-231, FM95, 22RV1, H2087) (**Figure 4a-d**). Next, we investigated the effect of this combination on tumor growth *in vivo* in a Class 2 BRAF mutant PDX colorectal cancer model (BVR-O-17), and two additional Class 2 BRAF mutant melanoma PDX models (GCRC-Mel1, GCRC-2015). B+E+P significantly inhibited tumor growth in these Class 2 BRAF mutant PDX models compared to vehicle treatment (**Figure 4e-g**). We observed inhibition of the MAPK pathway via ERK phosphorylation in all 3 PDX models treated with B+E. Palbociclib alone did not inhibit ERK phosphorylation but effectively inhibited RB phosphorylation. B+E+P inhibited both ERK and RB phosphorylation in the 3 models, though some phosphorylation of these proteins remained in certain tumors (**Figure 4h-j**). Similar results were observed in Class 3 BRAF mutant models (**Figure S7a-c**). We did not observe any increased toxicity with B+E+P compared to B+E, as evidence by no reduction in animals’ weights (**Figure S7d**). To further investigate the impact of B+E+P triple therapy on transcriptional outputs, we calculated the MAPK pathway activity score (MPAS) along with the E2F and G2M gene set scores from the RNA sequencing of these PDXs taken at the experimental end-point. B+E treatment significantly inhibited downstream MAPK pathway activity compared to vehicle-treated tumors; however, B+E did not impact transcript levels of E2F targets or G2M checkpoint gene sets. Conversely, treatment with Palbo alone inhibited expression of E2F targets and G2M checkpoints gene sets but did not impact MAPK pathway activity. However, B+E+P triple therapy significantly repressed all 3 gene sets (MPAS, E2F targets, and G2M checkpoint) compared to vehicle treatment in multiple PDX models (p<0.0001) (**Figure 4k-m; Figure S8**). These data reinforce the potential therapeutic benefit of combined MAPK pathway and CDK4/6 inhibition for the treatment of non-V600E *BRAF* mutant tumors.

**Figure 4:**
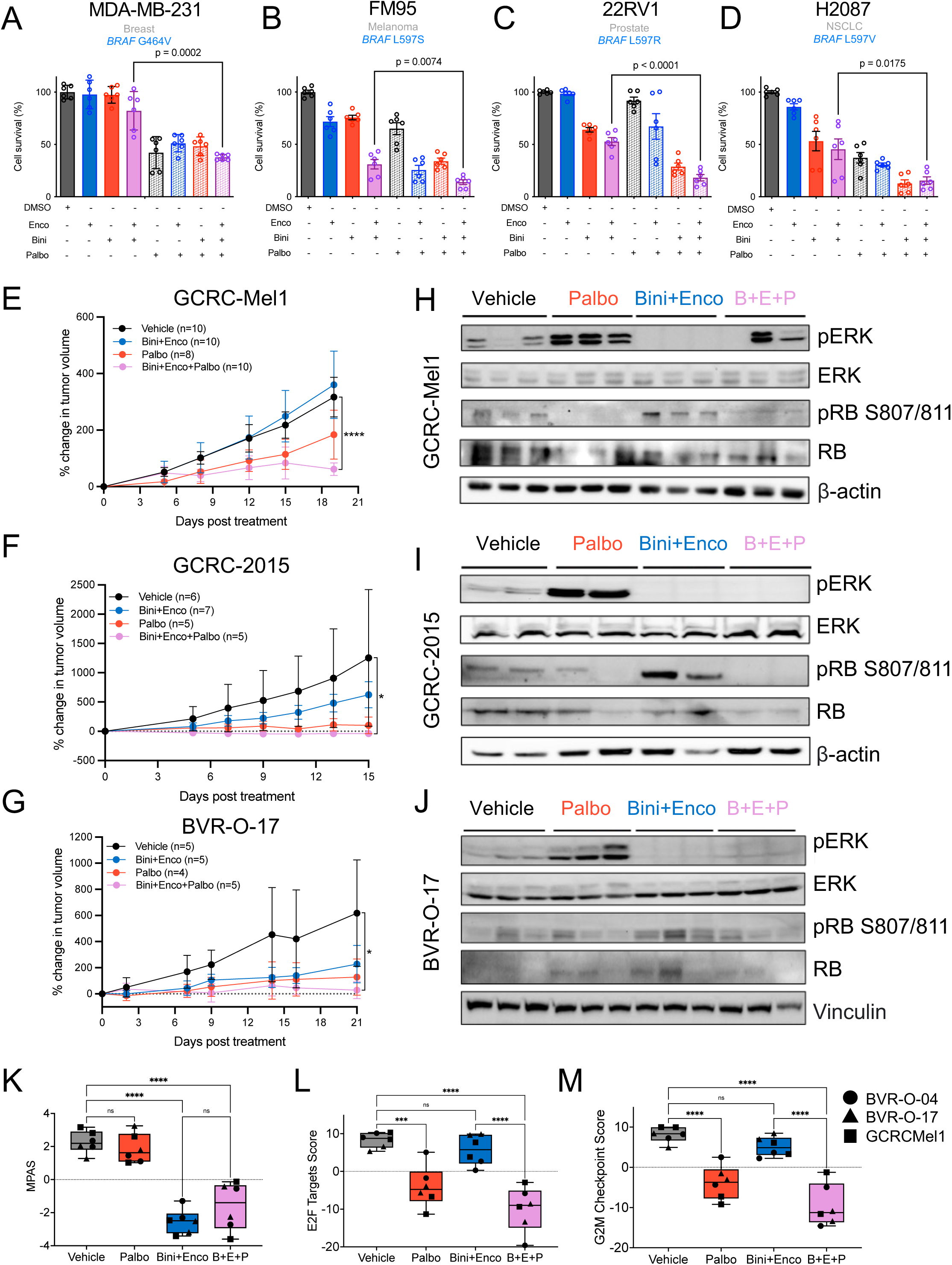
**Evaluation of CDK4/6 + BRAF/MEK inhibition in Class 2 BRAF mutant tumors** Quantification of clonogenic assays of cancer cells with endogenous Class 2 BRAF mutations that were treated with combinations of BRAF, MEK and CDK4/6 inhibitors, encorafenib, binimetinib and palbociclib respectively in **A)** HMV-II melanoma **B)** FM95 melanoma **C)** 22RV1 prostate cancer and **D)** H2087 NSCLC cells. Tumor growth curves of **E)** GCRC-Mel1, **F)** GCRC-2015, and **G)** BVR-O-17 PDXs treated with vehicle, palbociclib (80mg/kg/day) (palbo) for all PDXs, binimetinib (15mg/kg/d) + encorafenib (75mg/kg/d) (bini + enco) for GCRCMel1 and BVR-O-17, and binimetinib (20mg/kg/d) + encorafenib (75mg/kg/d) (bini + enco) for GCRC-2015. For the triple therapy of binimetinib + encorafenib + palbociclib (B+E+P), the doses of the respective drugs were 15/75/80 mg/kg/d for GCRCMel1, 20/75/80 mg/kg/d for GCRC2015, and 15/50/80 mg/kg/d for BVR-O-17. One vehicle-treated mouse (BVR-O-17) reached a humane end-point on Day 14. Corresponding immunoblots of tumors taken at experimental endpoint from mice bearing PDXs (**H)** GCRC-Mel1, **I)** GCRC-2015, and **J)** BVR-O-17 that were treated with vehicle, bini+enco, palbociclib, or bini+enco+palbo. Each lane represents protein lysate from a different biological replicate (tumor). **K)** MPAS calculated from the RNA expression of 10 genes comprising the MPAS signature from BVR-O-04 (circle), BVR-O-17 (triangle), and GCRC-Mel1 (square) treated with Vehicle, Bini+Enco, Palbo, or Bini+Enco+Palbo. **L)** E2F Targets score calculated from the RNA expression of 198 genes comprising the Hallmark MSigDB E2F Targets gene set from BVR-O-04 (circle), BVR-O-17 (triangle), and GCRC-Mel1 (square) treated with Vehicle, Bini+Enco, Palbo, or Bini+Enco+Palbo. **M)** G2M Checkpoint score calculated from the RNA expression of 190 genes comprising the Hallmark MSigDB E2F Targets gene set from BVR-O-04 (circle), BVR-O-17 (triangle), and GCRC-Mel1 (square) treated with Vehicle, Bini+Enco, Palbo, or Bini+Enco+Palbo. *P<0.05, **P<0.01, ***P<0.001, ****P<0.0001.

### Identifying SHP2 as a therapeutic target in BRAF/MEK inhibitor resistant Class 3 tumors

We sought to identify potentially actionable genes that are essential for the growth of Class 3 BRAF mutant tumors. To do this, we first mined the DepMap portal (23), (24) to define and compare the essential genes for growth of Class 3 (n=7) and Class 1 (n=117) BRAF mutant cancer cell lines(**Figure 5a**). We were interested in identifying genes that may be implicated in intrinsic B+E resistance in Class 3 BRAF mutant cancers. Multiple genes that were more essential in Class 3 cancer cells encoded proteins that were also constitutively activated in Class 3 BRAF mutant tumors from patients enrolled on the BEAVER trial. For example, 4/13 (31%) of patients with Class 3 BRAF mutations had co-occurring activating mutations in *RAF1* or *NRAS* (**Figure 1c**). We next investigated whether any of these essential genes were associated with acquired resistance to B+E in patients with Class 3 BRAF mutant cancer. We used the TSO500 targeted hybrid capture based next generation sequencing assay to quantify the mutations present in the ctDNA from two patients with Class 3 BRAF mutant cancer. Both patients initially experienced tumor regression with B+E treatment, but ultimately went on to develop disease progression (**Figure 5b**). Both patients had detectable ctDNA at baseline, but cleared their ctDNA after 1 cycle of B+E treatment. At the time of disease progression, one patient (BVR-O-07) developed a new *RAF1* mutation (p.R391W) that confers high kinase activity in a dimerization-dependent manner (25,26), and two new *MAP2K1* mutations that are both RAF-regulated, meaning that these *MAPK2K1* mutations require upstream RAF activity to function as oncogenes (27). A second patient (BVR-M-08) developed a new *TERT* promoter mutation and five new oncogenic *NRAS* mutations – each at different allele frequencies, suggestive of multiple subclones. We also identified a newly acquired *NRAS* mutation at the time of PD in a patient with Class 2 BRAF mutant melanoma who initially experienced a partial response (BVR-M-05; **Figure S9a**). In contrast, we did not observe any new acquired MAPK mutations in patients with Class 3 BRAF mutant cancer who experienced PD as best response, although one patient (BVR- O-13) did acquire a new loss of function mutation in the RB tumor suppressor at the time of PD (**Figure S9b**). Altogether, these somatic activating mutations in the MAPK pathway suggest a MAPK/ERK pathway-dependent mode of resistance that requires RAF-dimerization, which is a SHP2/RAS-dependent process.

**Figure 5:**
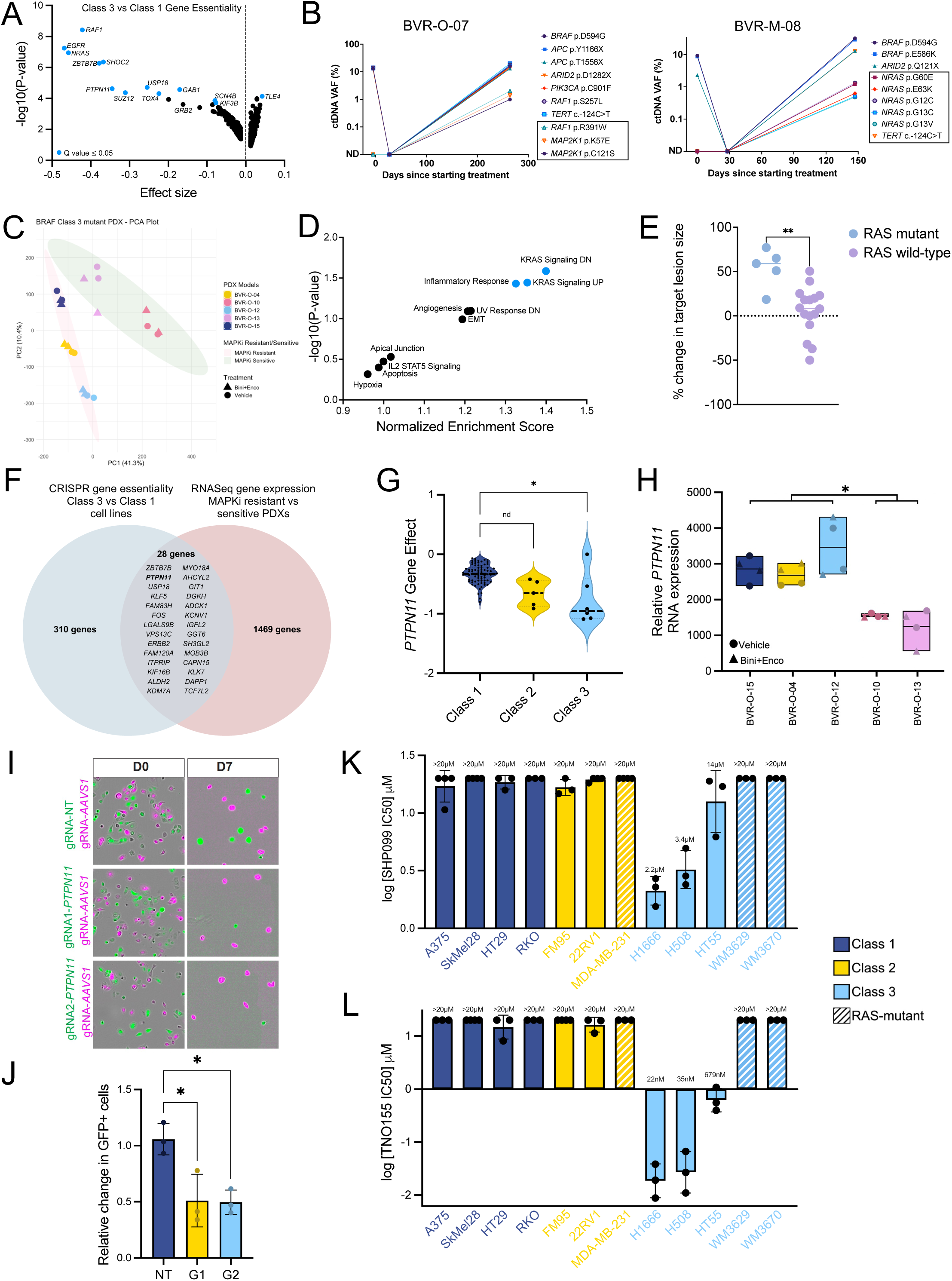
Identification of SHP2 as a therapeutic target in Class 3 BRAF mutant tumors. **A)** Dot plot analysis of Class 3 (n=7) vs Class 1 (n=117) BRAF mutant cancer cell lines with gene effect size (x-axis) plotted against -log10(P-value). Low effect size suggests gene essentiality in Class 3 vs Class 1 BRAF mutant cancer cell lines. **B)** Two patients (BVR-O-07 and BVR-M-05) with Class 3 BRAF mutant cancer experienced tumor regression with Bini+Enco treatment. Results of ctDNA analysis from samples taken prior to treatment, after completing 1 cycle (28 days) treatment and at progression is shown. Genes highlighted in boxes indicate mutations that were detected in ctDNA taken at time of progression that were not detected (ND) prior to starting treatment. VAF = variant allele frequency. **C)** Principal Component Analysis plot of the Bini+Enco and Vehicle treated Patient Derived Xenografts with Class 3 *BRAF* mutations (BVR-O-04/O-10/O-12/O-15/O-17). **D)** Normalized Enrichment Score from GSEA comparing Bini+Enco and Vehicle treated MAPKi Sensitive vs. MAPKi Resistant PDXs with Class 3 *BRAF* mutations (BVR-O-04/O-10/O-12/O-15/O-17). Plotted Gene Sets from the top 10 most enriched common gene sets between the 5 PDXs. **E)** Dot plot presenting the percentage change from baseline in target lesion size. Each dot represents a patient enrolled in the BEAVER trial with a RAS mutant (n=5) or RAS wild-type tumor (n=16). *RAS* mutation status was determined based on NGS of tumor tissue from archival or pre-treatment biopsies. **F)** Venn diagram showing unique and common genes between the CRISPR screen data from the DepMap in **A)** and significantly overexpressed genes in MAPKi-resistant PDXs vs their sensitive counterparts. **G)** Violin plot shows *PTPN11* gene essentiality in Class 1 (n=117), Class 2 (n=15), and Class 3 (n=7) BRAF mutant cell lines from the DepMap genome-wide CRISPR-Cas9 essentiality screen. **H)** Relative *PTPN11* gene expression in Bini+Enco and Vehicle treated MAPKi Sensitive vs. MAPKi Resistant PDXs. **I)** Representative images showing either H1666 cells expressing *PTPN11*-gRNA or NT-gRNA (both in green) co-cultured in a 1:1 ratio independently with H1666 cells expressing *AAVS1*-gRNA in magenta, at Day 0 and Day 7. **J)** Relative change in percentage of green population over total population was monitored over 7 days. **K)** SHP099 and **L)** TNO155 IC50 values across BRAF mutant cell lines, N=3 biological replicates plotted.

To identify additional genes that may be required for B+E-resistance, we investigated the transcriptomes of Class 3 BRAF mutant PDXs (n=5). Of these five PDXs, two were sensitive and three were intrinsically resistant to B+E (**Figure 2b, Figure S4c**). Principal component analysis of the PDX tumors revealed that the B+E sensitive tumors clustered together and away from the B+E-resistant tumors (**Figure 5c**). There were 3651 significantly differentially expressed genes between B+E-resistant vs. sensitive tumors (**Figure S10a-b**). The resistant tumors were significantly enriched for 3 gene sets, two of which were related to KRAS Signaling (**Figure 5d**). The enrichment of these gene sets supports the notion that RAS activation is involved in mediating resistance to B+E in the Class 3 BRAF mutant PDXs. Indeed, patients with activating *KRAS/NRAS* mutations experienced more tumor growth (relative increase in the sum of target lesion diameters) with B+E treatment compared to patients without *RAS* activating mutations at baseline (**Figure 5e)**. Of the 1469 overexpressed genes in resistant vs. sensitive Class 3 PDXs, 28 were included in the list of essential genes for Class 3 BRAF mutant cancers (**Figure 5f)**. Amongst these genes, *PTPN11* was one of the most essential genes in Class 3 cancer cell lines (**Figure 5g**) that was also more highly expressed in B+E-resistant Class 3 PDXs compared to B+E-sensitive PDXs (**Figure 5h**). *PTPN11* encodes the nonreceptor protein tyrosine phosphatase, SHP2. SHP2 activates the MAPK and PI3K pathways (28) through forming a complex with GRB2, GAB1, and SOS1 to activate the RAS superfamily of small GTPases (29), (30). In addition, SHP2 cooperates with several other proteins (EGFR, NRAS, CRAF) that were also found to be essential for the proliferation of Class 3 cancer cells (**Figure 5a**). In our analysis, we found that the two outlier Class 3 BRAF mutant cell lines with lower *PTPN11* gene essentiality scores (**Figure 5h**) harbor activating *RAS* mutations, hence the dispensability of *PTPN11* in these cell lines.

To confirm that *PTPN11* is an essential gene in Class 3 BRAF mutant cells, we performed a two-color CRISPR competition assay (**Figure 5i-j**). NCI-H1666 lung cancer cells stably expressing Cas9 were engineered to express either GFP in addition to a *PTPN11*-targeting gRNA, or mCherry in addition to a gRNA targeting the adeno-associated virus integration site (*AAVS1*) and were co-cultured together. Cells expressing 2 different *PTPN11*, but not non-targeting (NT), guide RNAs (gRNAs) were outcompeted by the *AAVS1* gRNA-expressing cells (i.e. wildtype for *PTPN11*), hence validating the predicted *PTPN11* gene essentiality for cell growth. Next, we assessed whether small molecule SHP2 inhibitors (SHP099, TNO155) could inhibit the growth of BRAF mutant cells *in vitro* (**Figure 5k-l**). As reported previously (7,31), we found that SHP2 inhibitors did not inhibit proliferation of Class 1 and 2 BRAF mutant cell lines. In contrast, while Class 3 BRAF mutant cells were sensitive to SHP2 inhibition, those with co-occuring *RAS* mutations (2 melanoma cell lines, WM3670, WM3629) were also insensitive to SHP2 inhibitor monotherapy. Together these results highlight SHP2 as a potential mediator of intrinsic and acquired resistance to B+E and a promising therapeutic target in Class 3 BRAF mutant cancers.

### Evaluating BRAF/MEK/SHP2 inhibitor combinations in Class 3 BRAF mutant cancers

We assessed whether the SHP2 inhibitor, TNO155, enhanced MAPK inhibitor-induced cell growth inhibition of Class 3 BRAF mutant cancer cells *in vitro*. The triple therapy combination (B+E+TNO155; B+E+T) was more effective at inhibiting cellular proliferation and inhibiting MAPK pathway activity vs. B+E and TNO (low dose of 100nM) in four Class 3 cell lines (H508, H1666, HT55, WM3670) (**Figure 6a**) and this corresponded with enhanced MAPK pathway inhibition by immunoblot in H508 and H1666 cells (**Figure 6b**). We did not observe any additional effect of adding SHP2 inhibitors to B+E in Class 2 BRAF mutant cancer cell lines (**Figure S11a-b**). Next, we investigated the effect of this drug combination on tumor growth *in vivo* using several Class 3 BRAF mutant PDX models. B+E+T significantly inhibited tumor growth or induced tumor regression in all PDX models tested, including those that were intrinsically resistant to B+E (BVR-O-12; BVR-O-15) (**Figure 6c-f**). However, this increased efficacy was also associated with increased weight loss with the B+E+T combination (**Figure S11c**). While we observed varying degrees of MAPK pathway inhibition in PDX models treated with TNO155 alone, B+E+T treatment consistently inhibited MAPK pathway (pERK) in all PDX models evaluated (**Figure 6g-i)**. Similarly, we calculated the MPAS and found that B+E+T treatment significantly inhibited downstream MAPK pathway activity compared to vehicle-treated tumors, more so than those treated with B+E treatment (**Figure 6j**). Altogether, these data support the potential benefit of using Shp2 inhibitors for the treatment of Class 3 BRAF mutant tumors to overcome resistance to BRAF/MEK inhibition.

**Figure 6:**
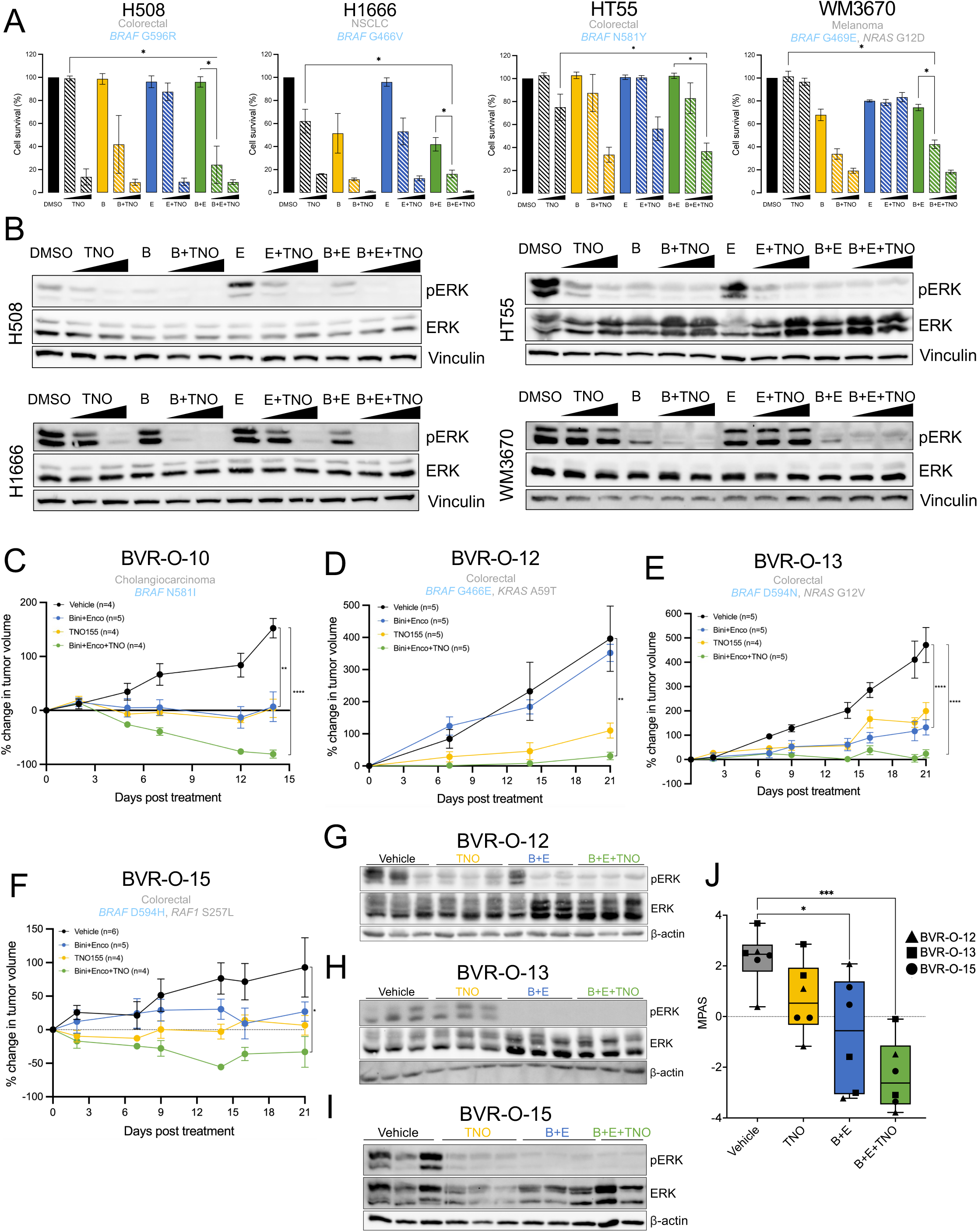
Evaluation of SHP2 + BRAF/MEK inhibition in Class 3 BRAF mutant tumors. **A)** Quantification of clonogenic assays of cancer cells with endogenous Class 3 *BRAF* mutations that were treated with combinations of BRAF, MEK and SHP2 inhibitors, encorafenib, binimetinib and TNO155 respectively in H508 (colorectal), H1666 (NSCLC), HT55 (colorectal) and WM3670 (melanoma) cells. Drug doses: H508 and HT55 - Bini 80nM, Enco 400nM, TNO 100nM and 1µM; H1666 - Bini 40nM, Enco 200nM, TNO 100nM and 1µM; WM3670 - Bini 120nM, Enco 600nM, 100nM and 1µM. N=3 biological replicates plotted. One-way ANOVA on AUC *P<0.05, **P<0.01, ***P<0.001, ****P<0.0001. **B)** Corresponding immunoblots of conditions from **A)** after 2hrs treatment. Tumor growth curves of **C)** BVR-O-10, **D)** BVR-O-12, and **E)** BVR-O-13 PDXs treated with Vehicle, Binimetinib (15mg/kg/d) + Encorafenib (75mg/kg/d) (Bini+Enco), TNO155 (10mg/kg/d) (TNO), or Binimetinib (15mg/kg/d) + Encorafenib (75mg/kg/d) + TNO155 (10mg/kg/day) (B+E+TNO). **F)** BVR-O-15 PDX treated with Vehicle, Binimetinib (15mg/kg/d) + Encorafenib (50mg/kg/d) (Bini+Enco), TNO155 (10mg/kg/d) (TNO), or Binimetinib (15mg/kg/d) + Encorafenib (50mg/kg/d) + TNO155 (10mg/kg/day) (B+E+TNO). One B+E+T-treated mouse (BVR-O-15) reached a humane end- point on Day 20. One-way ANOVA on AUC with Tukey correction for multiple comparisons *P<0.05, **P<0.01, ***P<0.001, ****P<0.0001. Corresponding immunoblots of tumors taken at experimental endpoint from mice bearing PDXs **(G)** BVR-O-12, **H)** BVR-O-13, and **I)** BVR-O-15 that were treated with Vehicle, Bini+Enco, TNO155, or Bini+Enco+TNO. Each lane represents protein lysate from a different biological replicate (tumor). **J)** MPAS calculated from the RNA expression of 10 genes comprising the MPAS signature from BVR-O-12 (triangle), BVR-O-13 (square), and BVR-O-15 (circle) treated with Vehicle, Bini+Enco, TNO, or Bini+Enco+TNO.

## Discussion

Here we describe the results of the investigator-initiated, Phase II BEAVER trial that evaluated the efficacy of BRAF/MEK inhibition in patients with advanced cancer with non-V600E *BRAF* mutations. We observed only minimal clinical efficacy of this regimen in a mixed patient population with a wide array of metastatic tumor types. This study did not meet the pre-specified criteria required for further investigation of this regimen in this patient population. We did, however, identify clinical, genomic, and transcriptomic characteristics that are associated with response and resistance to B+E, and these data could be useful in identifying specific sub- populations wherein this regimen may be more effective.

For example, we found that 3/5 patients with Class 2 and 3 BRAF mutant melanoma experienced tumor regression with B+E. While our study enrolled a small cohort of melanoma patients, this observation is supported by a separate clinical trial wherein 75% (3 out of 4) patients with Class 2 melanoma experienced a PR with low dose dabrafenib + trametinib (32). These clinical observations were reinforced by our *in vitro* experiments demonstrating that Class 2 BRAF mutant melanoma are more MAPK-dependent than non-melanoma tumor types. B+E induced more apoptosis in melanoma vs. non-melanoma cells and more potently repressed E2F and G2M gene sets in melanoma vs. non-melanoma cells. Enhanced MAPK pathway activity in the presence of B+E was a common feature of B+E-resistant melanoma but not of non-melanoma cells. Amongst melanoma patients who initially experienced tumor regression with B+E (BVR- M-05; BVR-M-08), newly acquired MAPK activating mutations in *NRAS* were observed at the time of disease progression, highlighting the MAPK-dependence of these tumors. Together, these data suggest that melanomas with Class 2 or 3 BRAF mutations may be more MAPK-dependent than other tumor types, and that this MAPK pathway-dependence may confer increased sensitivity to B+E. However, even amongst responders with melanoma, the duration of response or disease control with B+E was short and newly acquired resistance mutations developed quickly, highlighting the need for alternative therapeutic strategies to be developed.

The majority (86%) of patients did not respond to B+E treatment and we did not observe any newly acquired MAPK mutations at the time of progression in non-responders. These findings highlight that most tumors with Class 2 and 3 *BRAF* mutations may not be exquisitely dependent on MAPK pathway activity for tumor growth. Indeed, alterations in genes that regulate cell cycle progression were associated with intrinsic resistance to B+E and one patient, BVR-O-13, who experienced PD as best response, also developed a new loss-of-function mutation in the tumor suppressor RB1 at the time of progression. Moreover, p53 is a critical regulator of cell-cycle arrest in the context of cellular stress and DNA damage (33) and *TP53* mutations have been implicated in mediating resistance to targeted therapy (34). *TP53* was mutated in nearly 40% of tumors from patients on the BEAVER trial and all patients with *TP53* mutations experienced PD as best response, whereas none of the patients who experienced PR had a *TP53* mutation. Together, these data suggest that co-occurring alterations in cell cycle genes may uncouple the MAPK pathway from cell cycle regulation and nominate cell cycle mediators as potential therapeutic targets in these tumors.

We showed that CDK4/6 inhibitors could be combined with BRAF/MEK inhibitors, and this led to more cell growth inhibition *in vitro* and tumor growth inhibition *in vivo* in Class 2 and 3 BRAF mutant tumors. This therapeutic strategy may be particularly relevant in tumors where cell cycle progression is not uniquely regulated by the MAPK pathway. For example, in tumors where alterations in cell cycle regulators are present or in tumors with co-occurring MAPK-independent oncogenic alterations. These include RAS-PI3K-AKT pathway alterations such as those present in the BVR-O-04, BVR-O-13 and BVR-O-17 PDXs that were more responsive to B+E+P triple therapy vs. B+E alone. The combination of BRAF/MEK/CDK4/6 inhibition has previously been reported to be more effective than BRAF/MEK inhibition alone in preclinical models of Class 1 BRAF mutant melanoma, in part by modulating the tumor immune microenvironment (35). Currently, there is one on-going clinical trial evaluating the combination of binimetinib, encorafenib and palbociclib in patients with BRAF V600 mutant metastatic melanoma (NCT04720768). Our preclinical data support the clinical investigation of this regimen for Class 2 & 3 BRAF mutant cancers.

Several SHP2 inhibitors, including TNO155 are actively being investigated in clinical trials (NCT03114319, NCT03634982) for various types of cancers. We found that SHP2 inhibitors - when combined with BRAF/MEK inhibitors - could potentiate tumor regression and delay resistance, even in Class 3 BRAF mutant PDX models that were intrinsically resistant to B+E. Indeed, B+E+T led to a more profound inhibition of MAPK activity in Class 3 tumors compared to B+E treatment alone. It has been reported that some *RAS* mutations render tumors resistant to SHP2 inhibitors, however other KRAS mutant tumors are sensitive to Shp2 inhibitors ((36), (37), (7), (38)). Indeed, we found that multiple RAS co-mutated Class 3 BRAF mutant cancer cells were non-responsive to the SHP2 inhibitor monotherapy. Interestingly however, the addition of MAPK inhibitors potentiated responsiveness to TNO155 in *NRAS* G12D mutant WM3670 melanoma cells and in *NRAS* G12V or *KRAS* A59T co-mutated colorectal cancer PDXs. These findings suggest that SHP2i/MAPKi combinations may be effective even in RAS co-mutated Class 3 BRAF mutant tumors. Two patients with Class 3 *BRAF* mutations initially experienced tumor regression accompanied by ctDNA clearance, but developed new MAPK mutations at the time of disease progression. BVR-O-07 developed a new *RAF1* mutation and two new *MAP2K1* mutations that all require RAF-dimerization, which is a SHP2/RAS-dependent process. A second patient with a Class 3 BRAF mutation developed 5 new *NRAS* mutations at the time of progression. We have shown that SHP2i/MAPKi combinations remain effective even in NRAS co-mutated models of Class 3 BRAF mutant cancer. Thus, theoretically all these newly acquired MAPK-activating mutations would remain sensitive to SHP2i/MAPKi combinations, strengthening the rationale for pursuing this therapeutic strategy in patients with Class 3 BRAF mutant cancer. One note of caution, however, is that this triple therapy was associated with more weight loss in mice compared to B+E alone, and thus alternative dosing schedules or alternative SHP2 inhibitors may be required to mitigate enhanced toxicity.

Limitations of this study include the relatively small sample size and the fact that we did not complete enrollment of the BEAVER trial due to poor accrual. We cannot make any definitive claims about the efficacy of this regimen within specific cancer types because our sample size was too small.

The genomic complexity of Class 2 and 3 BRAF mutant tumors, relative to Class 1 BRAF mutant tumors, remains an important therapeutic challenge. Our findings suggest that many Class 2 and 3 BRAF mutant cancers can readily develop MAPK-dependent and MAPK-independent mechanisms of therapeutic resistance. Together, these data demonstrate that MAPK inhibition alone - even with novel and emerging next-generation MAPK inhibitors - may not yield deep and sustained therapeutic responses in these tumors. Future clinical trials aimed at developing precision therapies for Class 2 and 3 non-V600 BRAF mutations should incorporate inhibitors of proteins that regulate additional pathways beyond the MAPK pathway. Our data highlight CDK4/6 and SHP2 as viable therapeutic targets for future drug development strategies for these tumors.

## Materials & Methods

### BEAVER Trial Study Design

The BEAVER trial (Binimetinib and Encorafenib for the Treatment of Advanced Solid Tumors With Non-V600E *BRAF* Mutations, <u>NCT03839342</u>) enrolled patients from June 2019 to November 2023. It was approved by the University Health Network research ethics board (REB ID: 18-6324).

Key eligibility criteria were: patients with advanced solid tumors with non-V600E activating (Class 1 and 2) or inhibitory (Class 3) *BRAF* mutations, and no prior BRAF/MEK inhibitors. This was a single arm, open-label study. All patients received binimetinib (45mg PO BID) and encorafenib (450mg PO daily) on a 28-day cycle until intolerable toxicity or progression.

The primary objective was to evaluate the objective response rate (ORR) as per RECIST 1.1 criteria (39). Secondary objectives were to evaluate: progression free survival (PFS), overall survival (OS) and disease control rate (DCR). Exploratory objectives were to: 1) evaluate the dynamic changes and molecular evolution of circulating tumor DNA (ctDNA) profiles, before during and after treatment with binimetinib and encorafenib 2) establish patient-derived xenograft (PDX) models of advanced solid tumors with non-V600E *BRAF* mutations 3) evaluate biomarkers of response, and to identify molecular mechanisms of resistance to binimetinib and encorafenib in tumors with non-V600E *BRAF* mutations.

The BEAVER trial was a Simon 2-stage trial with the following statistical parameters: P0 = 0.05, P1 = 0.25, Alpha = 0.05, Power = 0.80. Seven patients were planned to be enrolled in the first stage. If 1 of 7 patients enrolled in the first stage achieved an objective response, the trial would advance to the second stage. In the second stage, up to 19 patients will be enrolled. If 4/26 patients enrolled in the entire study population achieved an objective response, the study drugs would be considered worthy of further evaluation. Additional details are provided in the Clinical Trial Protocol. The clinical trial schematic in Figure 1 was created using Biorender.com

### Statistical analyses of clinical data

Differences in objective responses according to clinical and genomic variables were assessed using Fisher’s exact test. Differences in tumor measurements according to genomic variables were assessed using an unpaired T-test. PFS and OS were visualized with a Kaplan-Meier curve and differences were assessed with a log-rank test. Statistical analyses were performed using Stata/MP v17.0.

### Sequencing of patients’ tumors and ctDNA

All patients enrolled on the BEAVER trial provided tumor tissue from archival specimens or from biopsies of metastatic tumors obtained prior to treatment initiation. Fresh tumor biopsies samples were collected with an 18-gauge core needle using standard surgical techniques. FFPE tissue. Genomic DNA and RNA were co-isolated from FFPE using standardized procedures with the Maxwell RSC RNA FFPE kit (Promega) in the Advanced Molecular Diagnostics Laboratory (AMDL) at the University Health Network in Toronto, ON. Sequencing of tumor tissue was performed using either the Oncomine Comprehensive Assay v3 (OCAv3) or the Illumina TruSight Oncology 500 (TSO500) assay. The OCAv3 and TSO500 assays evaluate 161 and 532 relevant cancer driver genes, respectively. For samples analyzed with the OCAv3 assay, sequencing was performed on the Ion S5 XL System and data analysis was performed using the Ion Reporter (ThermoFisher). Variant annotations were obtained from OncoKB. For samples analyzed with the TSO500 assay Sequencing was performed using the Illumina sequencing platform at the AMDL. Variant calls were generated using a custom bioinformatics pipeline with alignment to genome build GRCh37/hg19. Variant interpretation is based on results returned by the Qiagen QCI platform (v 7.1.20210428) along with searches of cancer variant databases and biomedical literature.

Cell-free (cfDNA) was extracted from blood plasma using MagMAX Cell-Free DNA Isolation kit and analyzed using the Illumina TSO500 ctDNA targeted hybrid capture based next generation sequencing assay. Sequencing was performed using the Illumina sequencing platform at AMDL. The coding regions and 5 bp of intronic regions were analyzed for variants. Variant calls were generated using the Illumina DRAGEN pipeline with alignment to genome build GRCh37/hg19. Variant interpretation is based on results returned by the Qiagen QCI platform (v 9.2.0.20230922) along with searches of cancer variant databases and biomedical literature. Minimal acceptable coverage for all reported variants was >800x. Variants of established, potential or uncertain clinical significance - considered Tier I, Tier II, and Tier III variants, respectively - were reported.

### Generation of PDX models

BVR PDX models were generated using fresh tissue from needle biopsies of metastatic tumors from patients enrolled on the BEAVER trial. Tissue fragments were implanted into the flanks of 3 female NSG mice. Mice were monitored for tumor development by caliper measurement. P0 tumors that grew were harvested and viable fragments were frozen or implanted as a P1 passage into female SCID mice. The development of GCRC PDXs was previously described (15).

### *In vivo* drug treatment experiments

For drug-treatment experiments with BVR PDXs, SCID mice were implanted with early passage (P1-P3) PDXs and monitored until the tumor volume reached ∼100mm^3^. Mice were then randomized to an indicated drug treatment regimen. All drug treatments were given by once or twice daily oral gavage. Drug treatment was held if BVR PDX-bearing mice lost >20% weight until they regained it. Tumor volume (*V*) was calculated as *V* = (length × width^2^)/2. The body weight of each mouse was recorded every 2 to 3 days. Mice were sacrificed at the days indicated or earlier if they reached a humane end-point. For each PDX model, Vehicle and Bini+Enco mice were reused in different tumor growth curves within the manuscript. Xenograft studies were designed and conducted following the institutional animal care guidelines, according to a protocol approved by the UHN Animal Care Committee. For GCRC PDXs, tumor fragments were explanted into cell culture and cells were counted. 1 million cells were implanted subcutaneously bilaterally (GCRC-Mel1) or unilaterally (GCRC-2015) into the flanks of female NSG mice. This study followed the institutional animal care guidelines, according to a protocol approved by the McGill Comparative Medicine and Animal Resources Centre. Treatment and tumor growth assessment was performed as described above for the BVR PDXs.

### Cell culture and generation of resistant cell lines

A375 (CRL-1619), SkMel28 (HTB-72), RKO (CRL-2577), HT29 (HTB-38), H2087 (CRL-5922), H1666 (CRL-5885), and H508 (CCL-253) were purchased from ATCC. HT55 (C919Q16) were purchased from Sigma. WM3629 (WM3629-01-0001) and WM3670 (WM3670-01-0001) were purchased from Rockland. MDA-MB-231, 22RV1, HMV-II, FM95 cells were from Dr. Peter Siegel. All cell lines were authenticated by whole-exome sequencing (Novogene). A375, H1666, H508, WM3629, FM95, HMV-II, H2087, and WM3670 cells were cultured in RPMI (Wisent, cat. no. 350-000CL) containing 10% FBS (5% FBS was used for H2087 cells) (Wisent, cat. no. 080-450) and 1% PS (Wisent, cat. no. 450201EL). SkMel28 and MDA-MB-231 cells were cultured in DMEM (Wisent, cat. no. 319-005-CL) containing 10% FBS (5% FBS was used for MDA-MB-231 cells) and 1% PS. RKO and HT55 cells were cultured in EMEM (Wisent, cat. no. 320-005-CL) containing 10% FBS and 1% PS. HT29 cells were cultured in McCoy’s 5A (Wisent, cat. no. 317-010-CL) containing 10% FBS and 1% PS. Resistant FM95 and HMV-II cells were cultured in media supplemented with 1% GlutaMAX (ThermoFisher, cat. no. 35050061). 22RV1, MDA-MB-231, and HMV-II binimetinib and encorafenib-resistant lines were generated by treating parental cells with increasing concentrations of binimetinib and encorafenib over a period of 8-12 months. The FM95 binimetinib and encorafenib resistant line was generated by seeding cells at low density and treating them with a high dose for approximately 2 months. Following colony formation, single cell clones were further seeded in a 96-well plate to be expanded. 22RV1 and FM95 resistant lines were maintained in 500 nM encorafenib and 100 nM binimetinib. MDA-MB-231 and HMV-II resistant cells were maintained in 1000 nM encorafenib and 200 nM binimetinib. Parental cells were also kept in culture during this period and treated with DMSO as control. Resistance was confirmed with binimetinib and encorafenib IC50 values at least 3 times greater than those of the corresponding parental cells. All cell lines used in this manuscript were tested for mycoplasma every 2 weeks by PCR (abm, cat. no. G238).

### Immunoblotting

Cells were lysed with 30-100uL of TNE lysis buffer [50 mM tris-HCl (pH 8.0), 150 mM NaCl, 1% NP-40, 2 mM EDTA, 250 mM sodium pyrophosphate dibasic, 100 mM β-Glycerophosphate disodium salt hydrate, cOmplete Mini Protease Inhibitor tablet (Sigma, cat. no. 11836153001)] was added to cells on ice. Flash-frozen tumor tissue samples (from experimental endpoint) were pulverized in liquid nitrogen using a mortar and pestle and were lysed on ice with 100-300uL of TNE lysis buffer. Following centrifugation, protein lysates were quantified using Bradford protein reagent (Bio-Rad, cat. no. 5000006). Equal amounts of protein were loaded on 4-12% SDS-PAGE gels and were transferred onto PVDF membranes (Bio-Rad, cat. no. 1620264) by semidry transfer (Bio-Rad Trans-Blot Turbo Transfer System). Membranes were blocked (1% BSA) and incubated with primary antibodies overnight at 4°C (see Supplemental Methods for antibodies and dilutions). Membranes were washed, incubated in secondary antibodies, and antibody detection was performed using Immobilon Forte Western HRP Substrate (cat. no. WBLUF0100). Bands were visualized using the Bio-Rad Chemi-Doc Imaging System or by X-ray films.

### DNA and RNA extraction

Parental and resistant cells were plated and the following day, treated for 24hrs with DMSO or the corresponding dose of Bini+Enco that the resistant cells are grown in. Flash-frozen tumor tissue samples were pulverized in liquid nitrogen using a mortar and pestle. Tumor tissue DNA and RNA was extracted with the Zymo Research Quick-DNA Microprep Kit (cat. no. D3020) and the Qiagen RNeasy Kit (cat. no. 74134), respectively, according to the manufacturer’s instructions. RNA quantification and quality assessment was performed using the NanoDrop Spectrophotometer ND-1000 (software version 3.8.1).

### RNA-sequencing and Analysis

Procedures to acquire RNA counts are described in supplemental methods. RNA counts were normalized using the DESeq2 (version 1.42.0) algorithm in R (version 4.3.2). The Wald test was used for p-value calculation and Benjamini-Hochberg false discovery rate (FDR) for the padj values. A baseMean cutoff of 50 was used for all heatmaps to filter out low counts. Gene Set Enrichment Analysis (GSEA) was performed on DESeq normalized counts with 1000 gene set permutations and comparing groups by Ratio of Classes metric.

### Clonogenic assays

Crystal violet assays were performed as previously described (15) by treating cells with the indicated concentrations of drug(s) for 7-10 days. Crystal violet stains were re-solubilized by adding 1mL of methanol to each well and incubating at room temperature for 1h on a rocking shaker. 100µL of the resolubilized crystal violet solution was transferred into a 96-well plate and the relative absorbance was measured at 570 nm using a plate reader (Perkin Elmer Enspire 2300). Assays were performed at least in triplicates.

### Functional Genomics

#### siRNA transfections

2.5 x 10^5^ MDA-MB-231 cells were seeded in 6 well-plates and transfected, the following day, with the indicated concentrations of non-targeting, or *CDK4*-targeting, or *CDK6*-targeting siRNA (siGENOME SMARTPool; Dharmacon) using lipofectamine 2000 (Thermo Fisher Scientific), as per the manufacturer’s protocol. The following day, cells were trypsinized and 5 x 10^3^ cells from each condition were seeded in 48-well plates and incubated at least for 6hrs before adding DMSO or Binimetinib + Encorafenib at the indicated concentrations. After adding the drug treatments, plates were immediately placed in the IncuCyte where the confluence of cells in each condition was monitored over 72hrs. In parallel to seeding cells for the proliferation assays, the remaining of siRNA-transfected cells were re-seeded and maintained in 6 well-plates for the same period (72hrs) to validate the siRNA efficiency at the experimental endpoint by immunoblotting.

#### Two-color CRISPR competition assay

This assay was performed as detailed previously (40) with some modifications. HEK293 FT cells were transfected with the LentiCas9-Blast vector (Addgene #52962) for lentiviral production. NCI-H1666 cells were then transduced with the produced viruses and selected with blasticidin (5 µg/ml) for 5 days to generate a stable Cas9-expressing cell line.

A non-targeting (NT) gRNA in addition to two gRNAs targeting *PTPN11* were cloned individually in Lentiguide-gRNA-NLS-GFP-2A-PURO plasmid (Addgene #185473; provided by Dr. Stephane Angers). The LentiGuide-puro-NLS-mCherry plasmid (Addgene #185474) expressing a gRNA targeting the safe harbor *AAVS1* locus was provided by Dr. Stephane Angers ((41)). HEK293-FT cells were transfected with these vectors independently to produce lentiviruses. NCI-H1666-Cas9 cells were then transduced with the produced viruses for 24hrs and left to recover for the two following days before selecting with puromycin (1.5 µg/ml) for 4 days. After selection, 75 x 10^3^ H1666-NT gRNA or H1666-*PTPN11* gRNA(s) were seeded with 75 x 10^3^ H1666-*AAVS1* gRNA in a 12 well-plate and incubated overnight. The following day, plates were imaged by the G/O/NIR optical module of IncuCyte S5 (Sartorius) to define the GFP-positive and mCherry-positive cell count/representation as a reference timepoint (day 0; D0). Two days later, cells were split 1:10 and propagated in a new 12 well-plate. Cells were split again (1:10) in the following days if they passed 50% confluence until they were imaged at day 7 (D7) as the endpoint.

## Data Availability

All data produced in the present study are available upon reasonable request to the authors

## Acknowledgements

The BEAVER clinical trial was sponsored by the Cancer Genomics Program of the Princess Margaret Cancer Centre. Funding for clinical operations of the BEAVER trial were provided by Pfizer. Exploratory objectives and preclinical experiments were funded by a Conquer Cancer Foundation Young Investigator Award and a Career Development Award to AANR, a Canadian Cancer Society Challenge Grant (#707457) to AANR and AS, a Canadian Institutes of Health Research (CIHR) Project Grant (#180379) to AANR and start-up funding from the TransMedTech Institute and Apogee Canada Research Excellence Fund and the Jewish General Hospital Foundation to AANR. Additional infrastructure support to carry out this research was provided by a John R. Evans Leaders Fund award (#42153) from the Canada Foundation for Innovation to AANR with matched funds from the Province of Quebec. AANR acknowledges salary support from a Fonds de Recherche du Québec – Santé (FRQS) Clinical Research Scholar Award; J.M. is the recipient of an Elizabeth Steffen Memorial Fellowship from McGill University Faculty of Medicine and Health Sciences and a FRQS doctoral award, ER is a recipient of a Marathon of Hope Data Science Award and a FRQS doctoral award, M.R. and I.S.B. are recipients of CIHR postdoctoral fellowships; IEE is a Peter Quinlan Fellow in Oncology from McGill University Faculty of Medicine and Health Sciences and a recipient of a FRQS postdoctoral fellowship. CM is a recipient of a CIHR Canada Graduate Scholarships-Masters (CGS-M) award and a Masters Research Training Award from the Canadian Cancer Society. The schematic in Figures 1a and 3c was created with Biorender.com. We are thankful to staff members of the Cancer Genomics Program who were involved with administration and coordination of the BEAVER trial including: Celeste Yu, Elizabeth Shah, Samanta del Rossi and Sam Felicen, as well as technical support from Peter Tai, Lucy An, and Dr. Huijie Wang for pre-clinical experiments. We are especially grateful to all of the patients (and their families) who volunteered to enroll in this study.

## Conflicts of interest

Dr. Rose has provided consultation for Advanced Accelerator Applications/Novartis, EMD Serrono and Pfizer. Dr. Rose reports research funding from AstraZeneca (Inst), Novartis (Inst), Merck (Inst), Seattle Genetics (Inst), Pfizer (Inst), and Essa Pharma (Inst).

Dr. Soria Bretones is an employee of Repare Therapeutics

Dr. King reports research funding from Pfizer (Inst) and is an employee of GeneDx, Gaithersburg MD.

Dr. Pugh has provided consultation for AstraZeneca, Chrysalis Biomedical Advisors, and Merck (compensated); and receives research support (institutional) from Roche/Genentech and AstraZeneca.

Dr. Shepherd has provided consultation for AstraZeneca.

Dr. Razak reported a consulting/advisory role with Adaptimmune, Bayer, GlaxoSmithKline, Medison, Inhibrx, research funding from Deciphera, Karyopharm Therapeutics, Pfizer, Roche/Genentech, Bristol Myers Squibb, MedImmune, Amgen, GlaxoSmithKline, Blueprint Medicines, Merck, AbbVie, Adaptimmune, Iterion Therapeutics, Neoleukin Therapeutics, Daiichi Sankyo, Symphogen, Rain Therapeutics, and expert testimony with Medison.

Dr. Hansen reported receiving research funds (paid to institution) from: Advancell, AVEO, BMS, Janssen, Macrogenics, MSD, Seagen, Roche and Tyra Biosciences. Consulting fees (paid personally) from: Astellas, Bayer, Eisai, MSD.

Dr. Bedard reported an uncompensated consulting/advisory role with BMS, Pfizer, Seattle Genetics, Lilly, Amgen, Merck, Gilead Sciences, Repare and research funding from Bristol Myers Squibb (Inst), Sanofi (Inst), AstraZeneca (Inst), Genentech/Roche (Inst), GlaxoSmithKline (Inst), Novartis (Inst), Nektar (Inst), Merck (Inst), Seattle Genetics (Inst), Immunomedics (Inst), Lilly (Inst), Amgen (Inst), Bicara Therapeutics (Inst), Zymeworks (Inst), Bayer (Inst), Medicenna (Inst), Day One Biopharmaceuticals (Inst).

Dr. Siu reported consultant/advisory roles for: Merck, Pfizer, AstraZeneca, Roche, GlaxoSmithKline, Voronoi, Arvinas, Navire, Relay, Marengo, Daiichi Sankyo, Amgen, Medicenna, LTZ Therapeutics, Tubulis, Marengo, Nerviano, Pangea, Incyte, Gilead; Institutions receives grant/research support for clinical trials from: Merck, Novartis, Bristol-Myers Squibb, Pfizer/Seattle Genetics, Boerhinger-Ingelheim, GlaxoSmithKline, Roche/Genentech, AstraZeneca/Medimmune, Bayer, Abbvie, Amgen, Symphogen, EMD Serono, 23Me, Daiichi Sankyo, Gilead, Marengo, Incyte, LegoChem, Loxo/Eli Lilly, Medicenna, Takara; Spouse has leadership position: Treadwell Therapeutics; Spouse has stock ownership: Agios.

Dr. Spreafico reported a consulting/advisory role with Merck, Bristol-Myers Squibb, and Alents andgrant/research funding from Novartis, Bristol-Myers Squibb, Symphogen AstraZeneca/Medimmune, Merck, Bayer, Surface Oncology, Northern Biologics, Janssen Oncology/Johnson & Johnson, Roche, Regeneron, Alkermes, Array Biopharma/Pfizer, GSK, NuBiyota, Oncorus, Treadwell, Amgen, ALX Oncology, Nubiyota, Genentech, Seagen, Servier, Incyte, Alentis.

All other co-authors report no conflicts of interest.

