## Supplemental Figures for "CDK4/6 and SHP2 mediate BRAF/MEK inhibitor resistance in Class 2 and 3 BRAF mutant cancers"

A

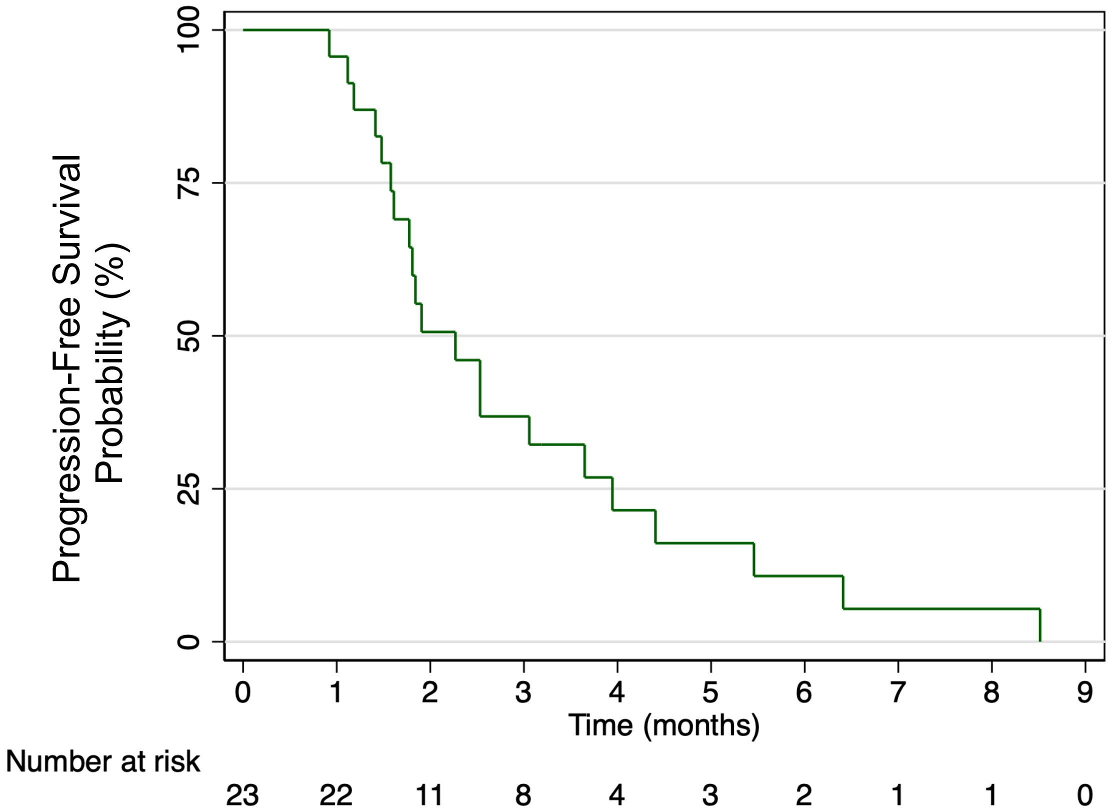

Progression-Free Survival Probability (%)
100
75
50
25
0
0 1 2 3 4 5 6 7 8 9
Time (months)
Number at risk
23 22 11 8 4 3 2 1 1 0


B

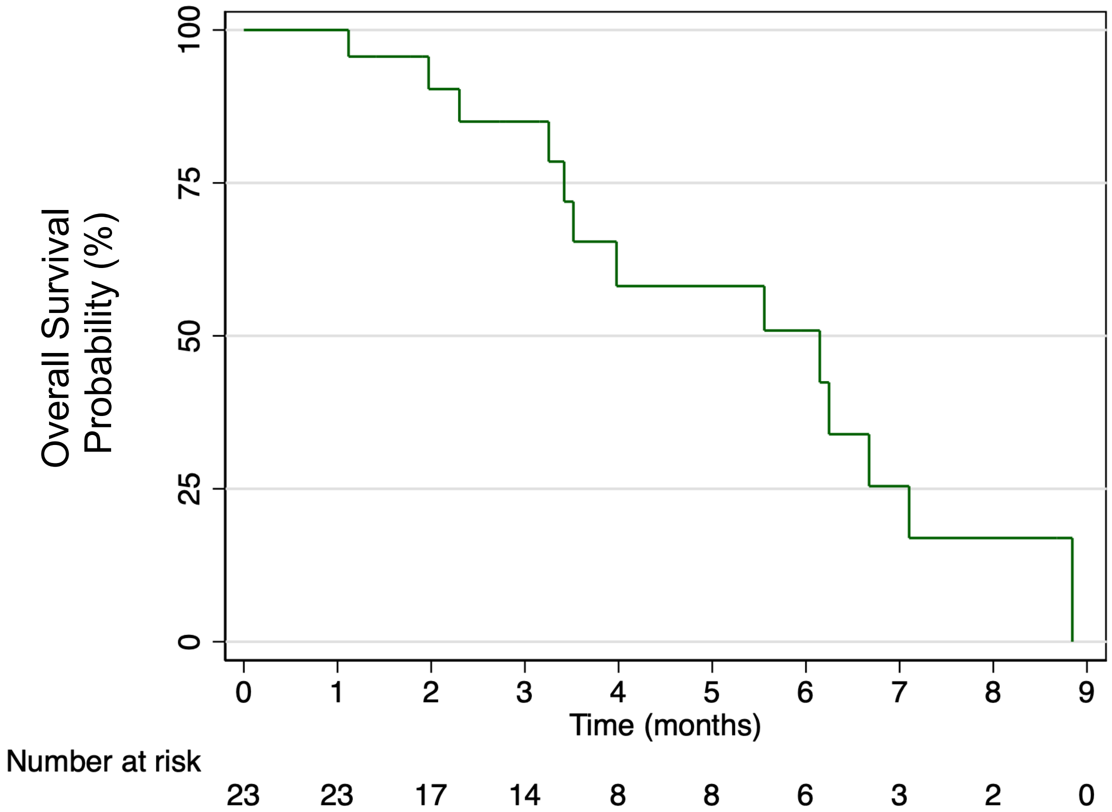

Overall Survival Probability (%)
100
75
50
25
0
0 1 2 3 4 5 6 7 8 9
Time (months)
Number at risk
23 23 17 14 8 8 6 3 2 0


**Figure S1**

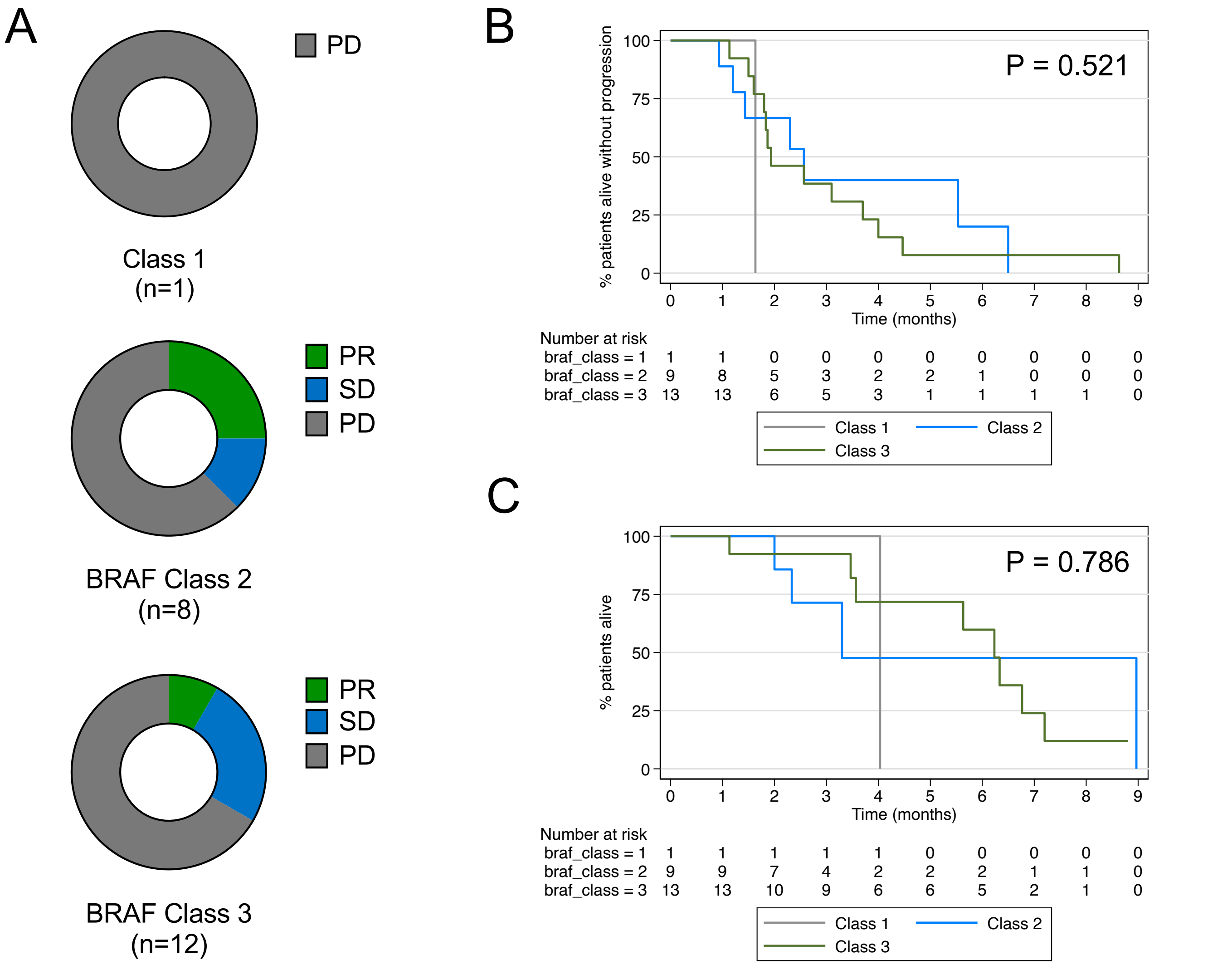
A
PD
Class 1
(n=1)
PR
SD
PD
BRAF Class 2
(n=8)
PR
SD
PD
BRAF Class 3
(n=12)
B
% patients alive without progression
P = 0.521
Time (months)
Number at risk
braf_class = 1 1 1 0 0 0 0 0 0 0 0
braf_class = 2 9 8 5 3 2 2 1 0 0 0
braf_class = 3 13 13 6 5 3 1 1 1 1 0
Class 1
Class 2
Class 3
C
% patients alive
P = 0.786
Time (months)
Number at risk
braf_class = 1 1 1 1 1 1 0 0 0 0 0
braf_class = 2 9 9 7 4 2 2 2 1 1 0
braf_class = 3 13 13 10 9 6 6 5 2 1 0
Class 1
Class 2
Class 3

**Figure S2**

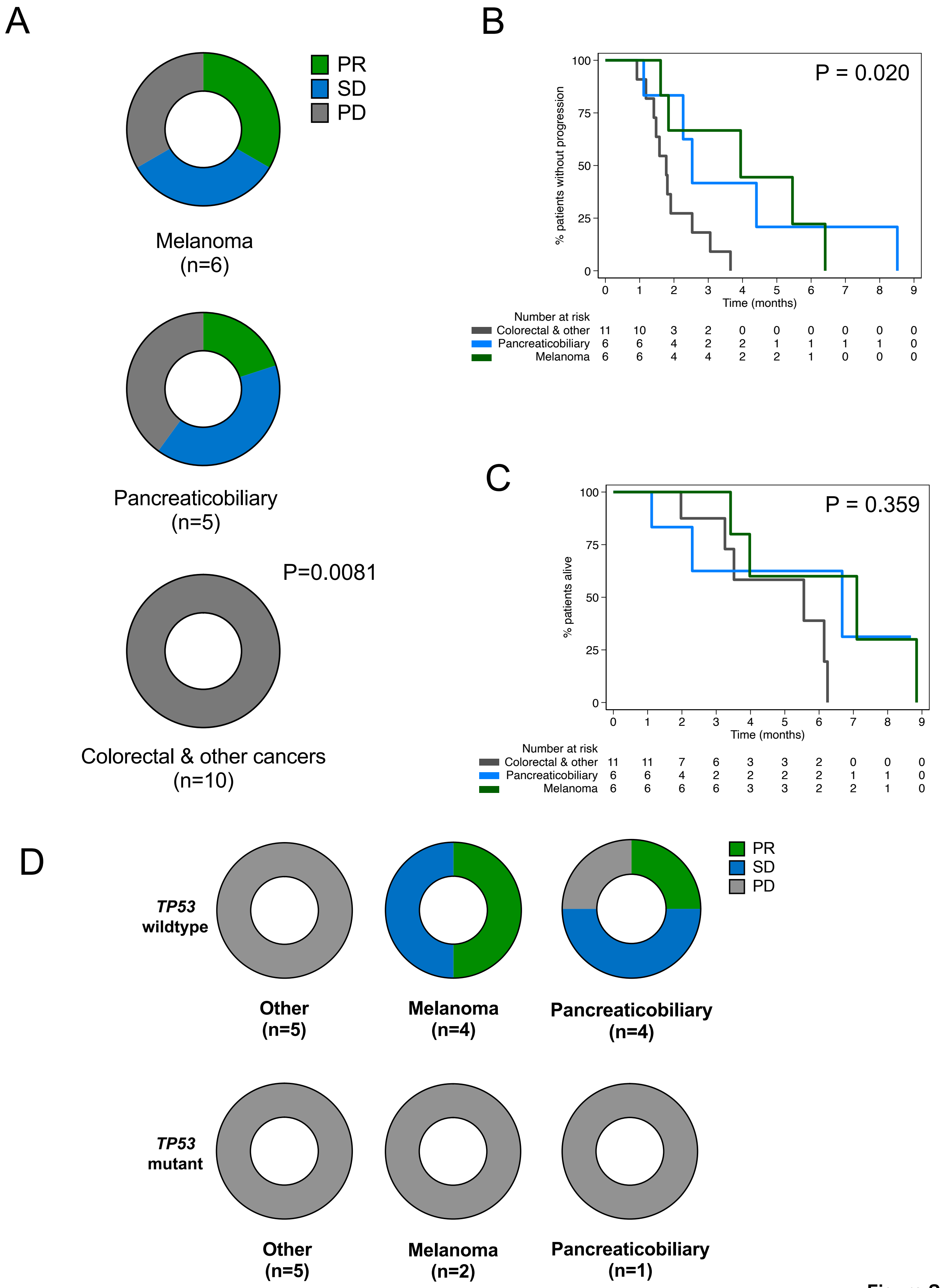
A
PR
SD
PD
Melanoma
(n=6)
Pancreaticobiliary
(n=5)
P=0.0081
Colorectal & other cancers
(n=10)
B
P = 0.020
% patients without progression
100
75
50
25
0
0 1 2 3 4 5 6 7 8 9
Time (months)
Number at risk
Colorectal & other 11 10 3 2 0 0 0 0 0 0
Pancreaticobiliary 6 6 4 2 2 1 1 1 1 0
Melanoma 6 6 4 4 2 2 1 0 0 0
C
P = 0.359
% patients alive
100
75
50
25
0
0 1 2 3 4 5 6 7 8 9
Time (months)
Number at risk
Colorectal & other 11 11 7 6 3 3 2 0 0 0
Pancreaticobiliary 6 6 4 2 2 2 2 1 1 0
Melanoma 6 6 6 6 3 3 2 2 1 0
D
TP53 wildtype
Other
(n=5)
Melanoma
(n=4)
Pancreaticobiliary
(n=4)
PR
SD
PD
TP53 mutant
Other
(n=5)
Melanoma
(n=2)
Pancreaticobiliary
(n=1)

**Figure S3**

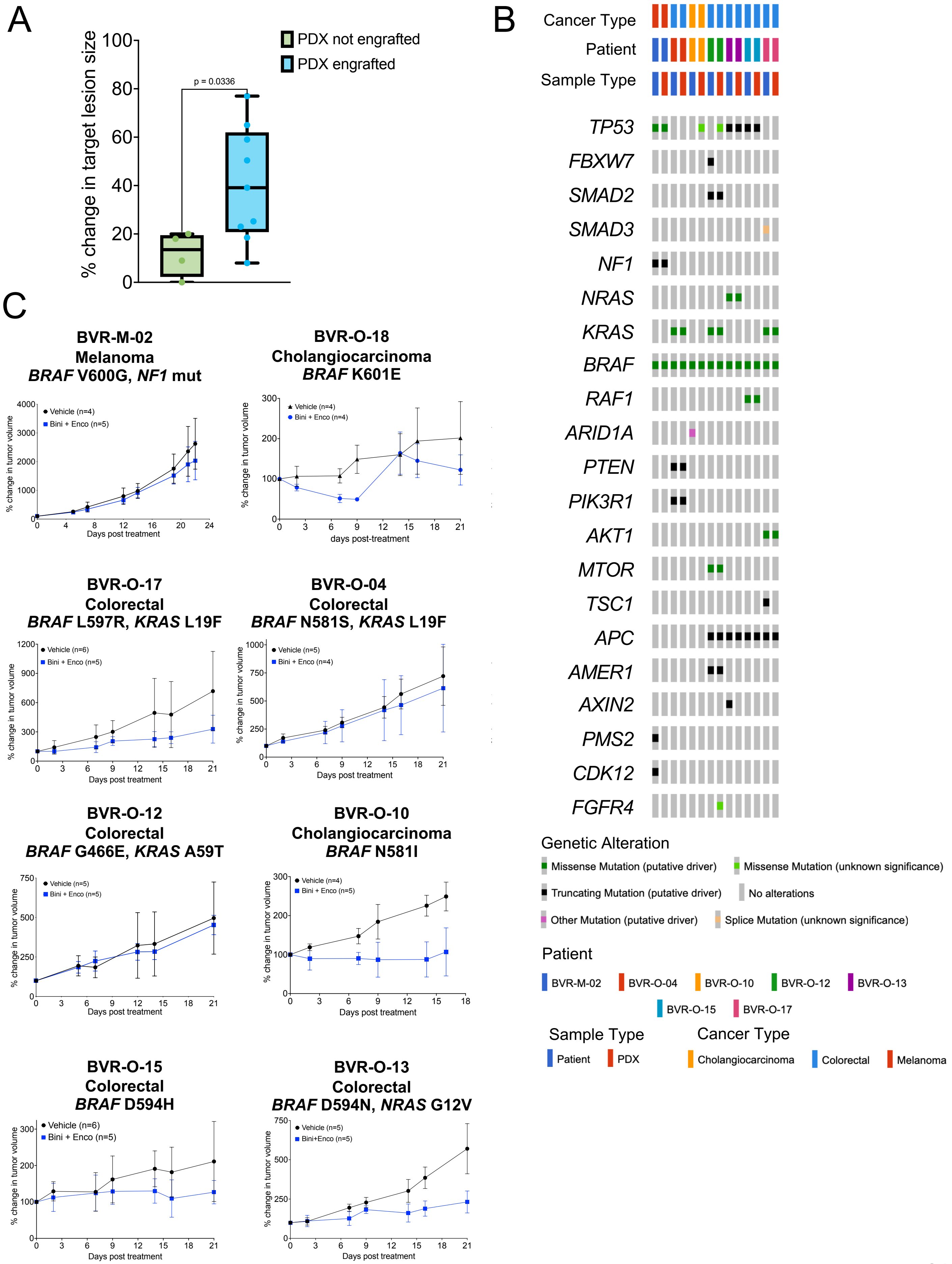
A
% change in target lesion size
PDX not engrafted
PDX engrafted
p = 0.0336
B
Cancer Type
Patient
Sample Type
TP53
FBXW7
SMAD2
SMAD3
NF1
NRAS
KRAS
BRAF
RAF1
ARID1A
PTEN
PIK3R1
AKT1
MTOR
TSC1
APC
AMER1
AXIN2
PMS2
CDK12
FGFR4
Genetic Alteration
Missense Mutation (putative driver)
Missense Mutation (unknown significance)
Truncating Mutation (putative driver)
No alterations
Other Mutation (putative driver)
Splice Mutation (unknown significance)
Patient
BVR-M-02
BVR-O-04
BVR-O-10
BVR-O-12
BVR-O-13
BVR-O-15
BVR-O-17
Sample Type
Patient
PDX
Cancer Type
Cholangiocarcinoma
Colorectal
Melanoma
C
BVR-M-02
Melanoma
BRAF V600G, NF1 mut
Vehicle (n=4)
Bini + Enco (n=5)
% change in tumor volume
Days post treatment
BVR-O-18
Cholangiocarcinoma
BRAF K601E
Vehicle (n=4)
Bini + Enco (n=4)
days post-treatment
BVR-O-17
Colorectal
BRAF L597R, KRAS L19F
Vehicle (n=6)
Bini + Enco (n=5)
BVR-O-04
Colorectal
BRAF N581S, KRAS L19F
Vehicle (n=5)
Bini + Enco (n=4)
BVR-O-12
Colorectal
BRAF G466E, KRAS A59T
Vehicle (n=5)
Bini + Enco (n=5)
BVR-O-10
Cholangiocarcinoma
BRAF N581I
Vehicle (n=4)
Bini + Enco (n=5)
BVR-O-15
Colorectal
BRAF D594H
Vehicle (n=6)
Bini + Enco (n=5)
BVR-O-13
Colorectal
BRAF D594N, NRAS G12V
Vehicle (n=5)
Bini+Enco (n=5)

**Figure S4**

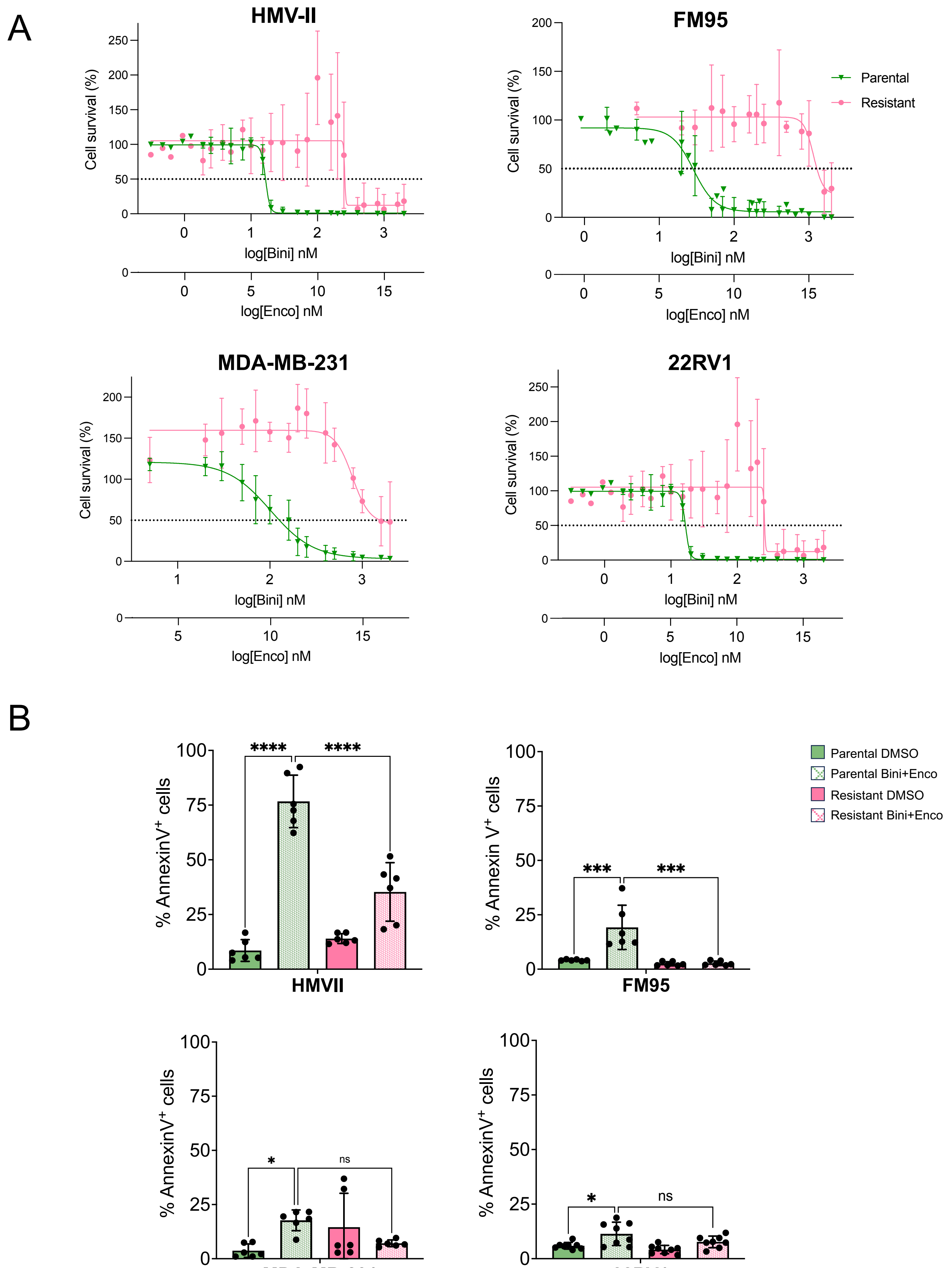
A
HMV-II
FM95
MDA-MB-231
22RV1
Cell survival (%)
log[Bini] nM
log[Enco] nM
Parental
Resistant
B
% AnnexinV+ cells
% Annexin V+ cells
HMVII
FM95
MDA-MB-231
22RV1
****
***
*
ns
Parental DMSO
Parental Bini+Enco
Resistant DMSO
Resistant Bini+Enco

Figure S5

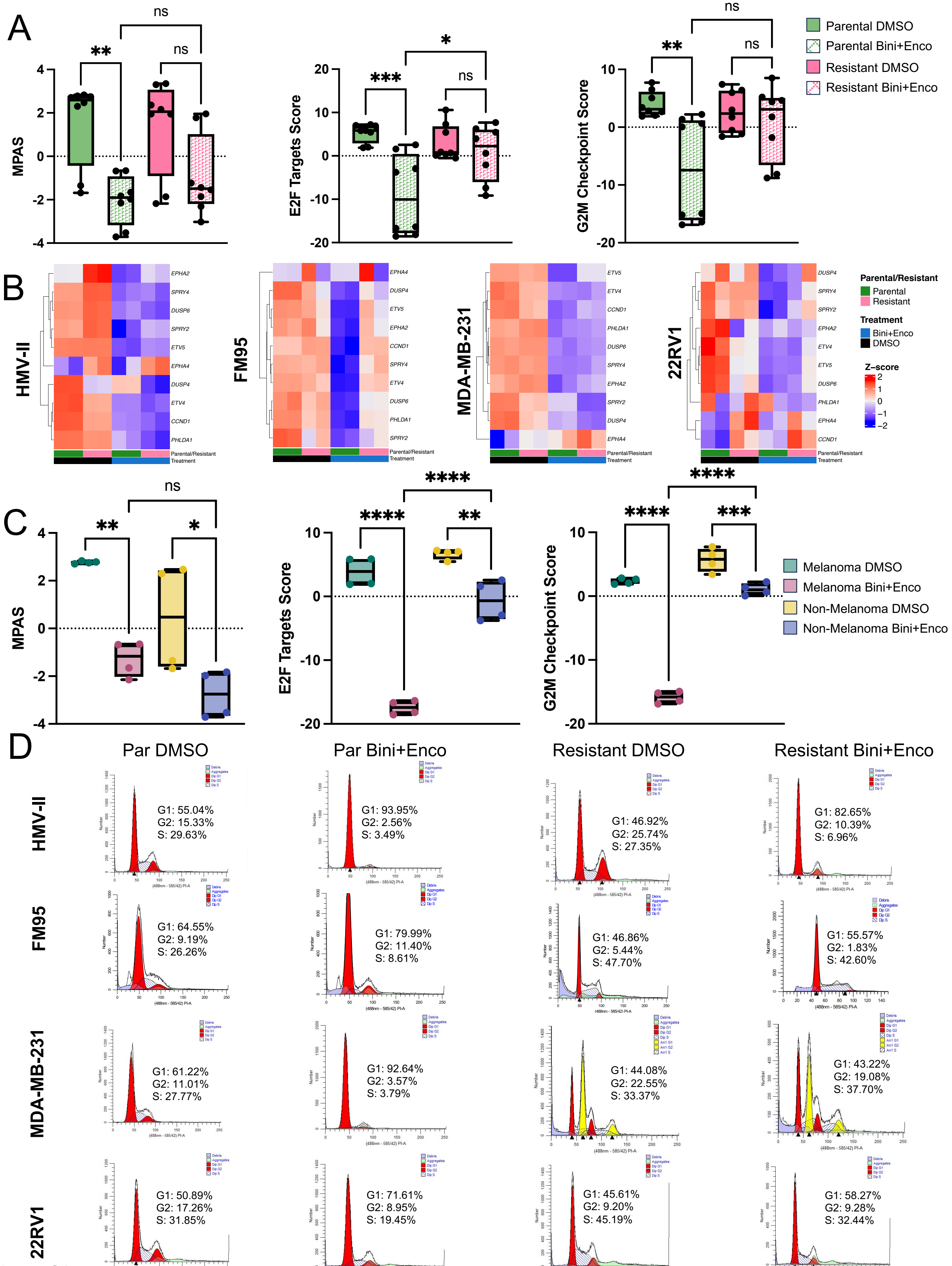
A
MPAS
E2F Targets Score
G2M Checkpoint Score
Parental DMSO
Parental Bini+Enco
Resistant DMSO
Resistant Bini+Enco
B
HMV-II
FM95
MDA-MB-231
22RV1
Parental/Resistant
Parental
Resistant
Treatment
Bini+Enco
DMSO
Z-score
C
Melanoma DMSO
Melanoma Bini+Enco
Non-Melanoma DMSO
Non-Melanoma Bini+Enco
D
Par DMSO
Par Bini+Enco
Resistant DMSO
Resistant Bini+Enco
G1: 55.04% G2: 15.33% S: 29.63%
G1: 93.95% G2: 2.56% S: 3.49%
G1: 46.92% G2: 25.74% S: 27.35%
G1: 82.65% G2: 10.39% S: 6.96%
G1: 64.55% G2: 9.19% S: 26.26%
G1: 79.99% G2: 11.40% S: 8.61%
G1: 46.86% G2: 5.44% S: 47.70%
G1: 55.57% G2: 1.83% S: 42.60%
G1: 61.22% G2: 11.01% S: 27.77%
G1: 92.64% G2: 3.57% S: 3.79%
G1: 44.08% G2: 22.55% S: 33.37%
G1: 43.22% G2: 19.08% S: 37.70%
G1: 50.89% G2: 17.26% S: 31.85%
G1: 71.61% G2: 8.95% S: 19.45%
G1: 45.61% G2: 9.20% S: 45.19%
G1: 58.27% G2: 9.28% S: 32.44%

Figure S6

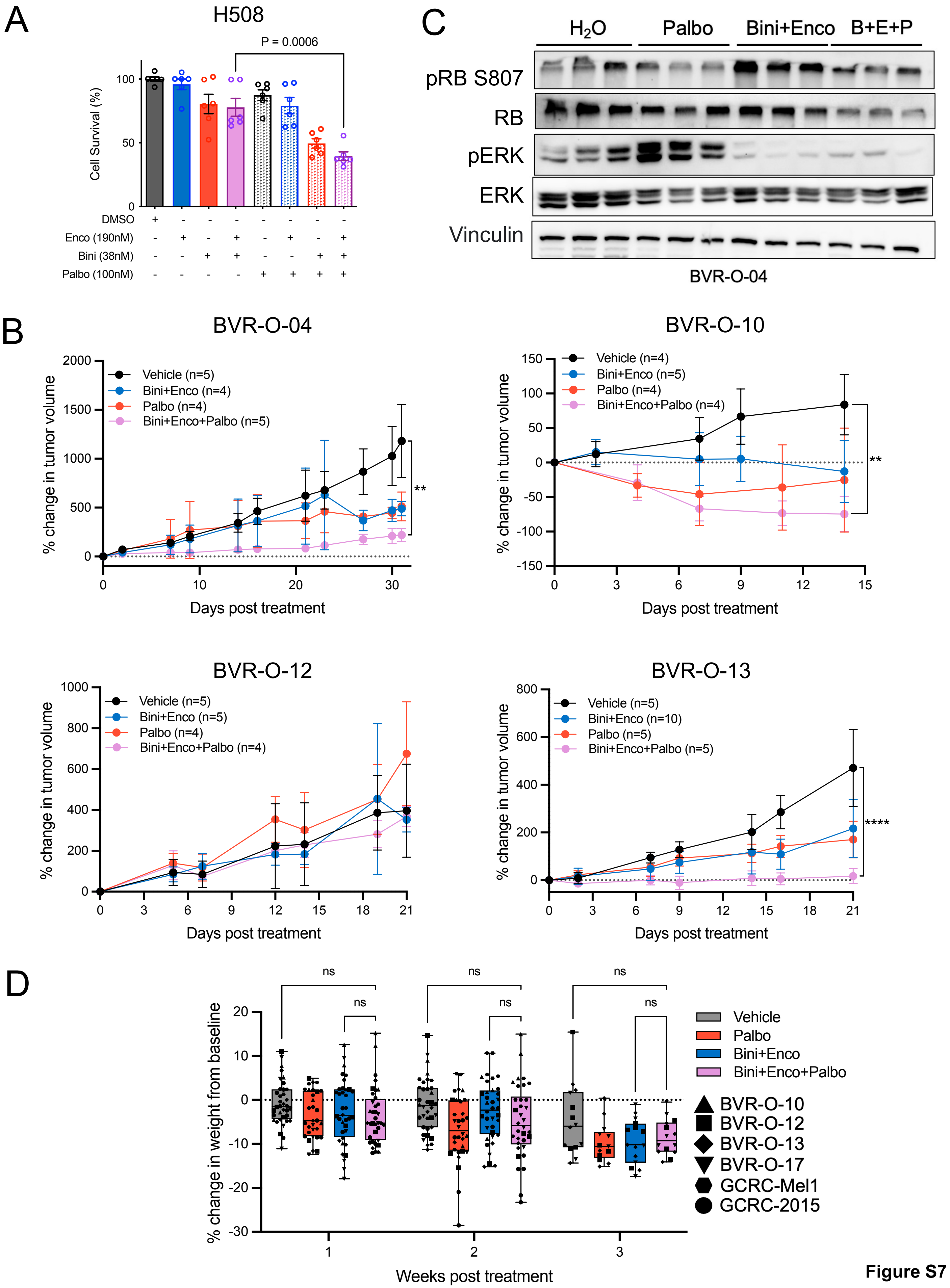
A
H508
P = 0.0006
Cell Survival (%)
100
50
0
DMSO
Enco (190nM)
Bini (38nM)
Palbo (100nM)
C
H2O
Palbo
Bini+Enco
B+E+P
pRB S807
RB
pERK
ERK
Vinculin
BVR-O-04
B
BVR-O-04
Vehicle (n=5)
Bini+Enco (n=4)
Palbo (n=4)
Bini+Enco+Palbo (n=5)
% change in tumor volume
Days post treatment
**
BVR-O-10
Vehicle (n=4)
Bini+Enco (n=5)
Palbo (n=4)
Bini+Enco+Palbo (n=4)
% change in tumor volume
Days post treatment
**
BVR-O-12
Vehicle (n=5)
Bini+Enco (n=5)
Palbo (n=4)
Bini+Enco+Palbo (n=4)
% change in tumor volume
Days post treatment
BVR-O-13
Vehicle (n=5)
Bini+Enco (n=10)
Palbo (n=5)
Bini+Enco+Palbo (n=5)
% change in tumor volume
Days post treatment
****
D
ns
% change in weight from baseline
Weeks post treatment
Vehicle
Palbo
Bini+Enco
Bini+Enco+Palbo
BVR-O-10
BVR-O-12
BVR-O-13
BVR-O-17
GCRC-Mel1
GCRC-2015

Figure S7

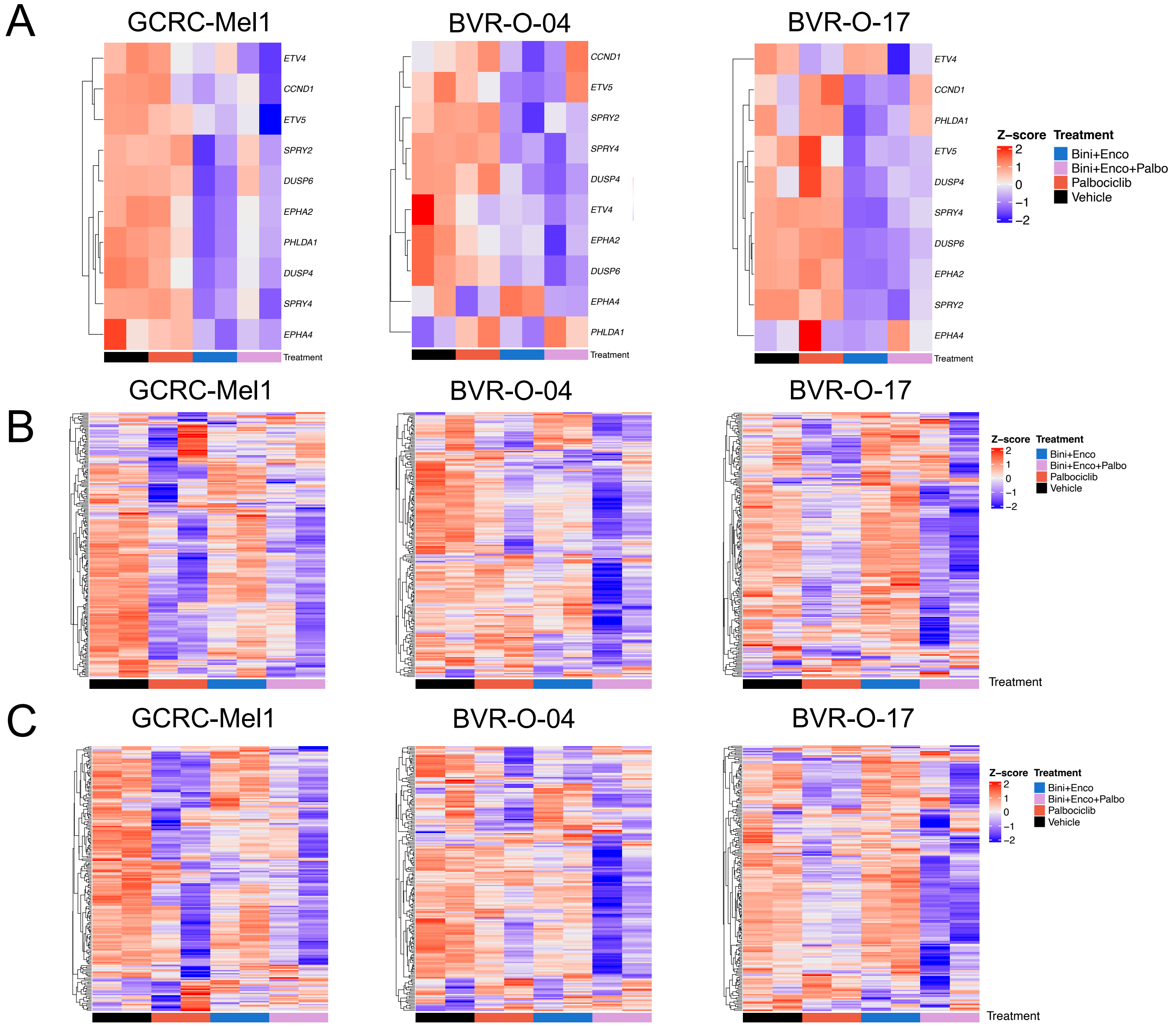
A
GCRC-Mel1
BVR-O-04
BVR-O-17
ETV4
CCND1
ETV5
SPRY2
DUSP6
EPHA2
PHLDA1
DUSP4
SPRY4
EPHA4
Treatment
Z-score
Bini+Enco
Bini+Enco+Palbo
Palbociclib
Vehicle
B
C

**Figure S8**

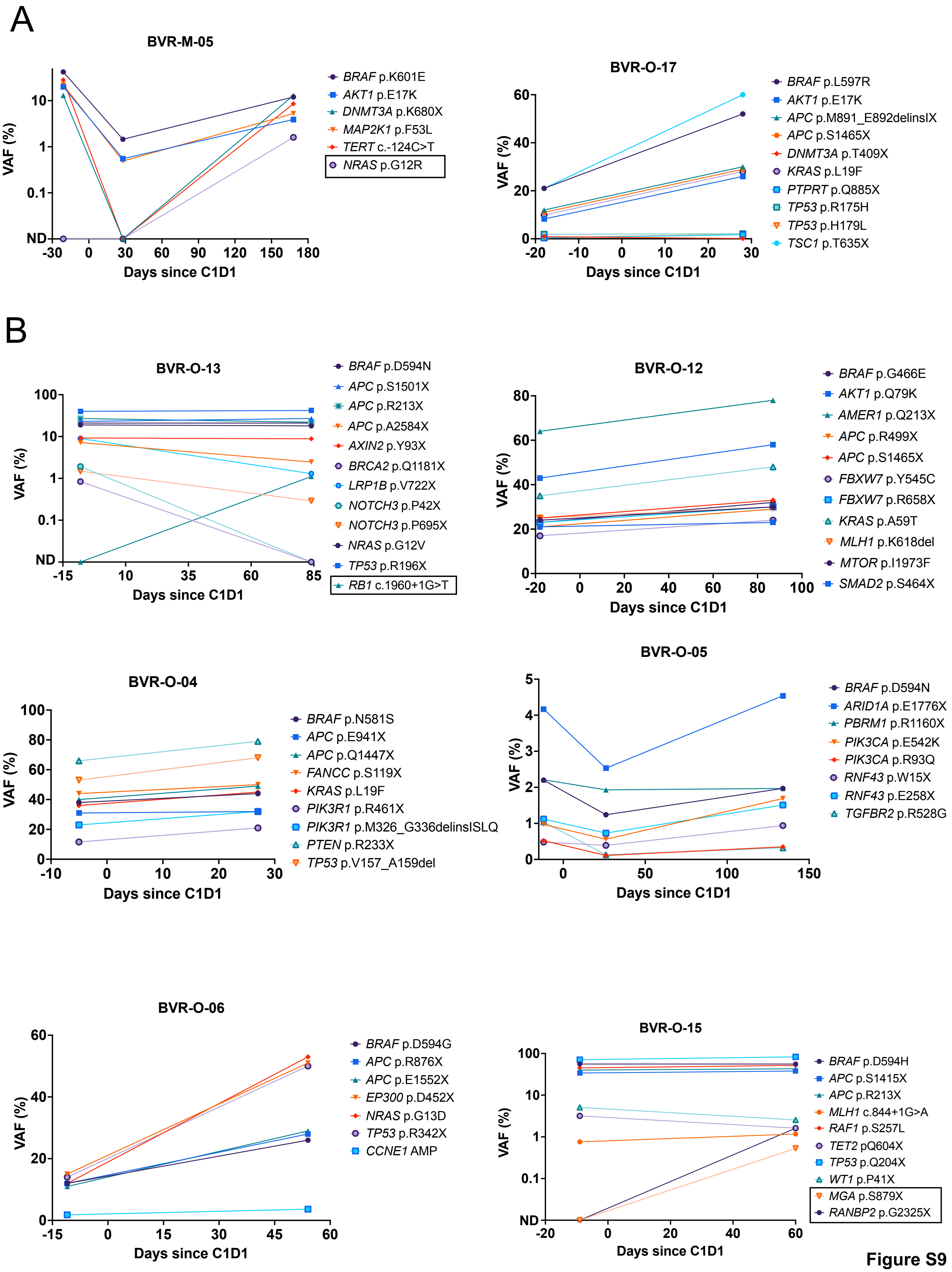
A
BVR-M-05
VAF (%)
Days since C1D1
BRAF p.K601E
AKT1 p.E17K
DNMT3A p.K680X
MAP2K1 p.F53L
TERT c.-124C>T
NRAS p.G12R
BVR-O-17
BRAF p.L597R
AKT1 p.E17K
APC p.M891_E892delinsIX
APC p.S1465X
DNMT3A p.T409X
KRAS p.L19F
PTPRT p.Q885X
TP53 p.R175H
TP53 p.H179L
TSC1 p.T635X
B
BVR-O-13
BRAF p.D594N
APC p.S1501X
APC p.R213X
APC p.A2584X
AXIN2 p.Y93X
BRCA2 p.Q1181X
LRP1B p.V722X
NOTCH3 p.P42X
NOTCH3 p.P695X
NRAS p.G12V
TP53 p.R196X
RB1 c.1960+1G>T
BVR-O-12
BRAF p.G466E
AKT1 p.Q79K
AMER1 p.Q213X
APC p.R499X
APC p.S1465X
FBXW7 p.Y545C
FBXW7 p.R658X
KRAS p.A59T
MLH1 p.K618del
MTOR p.I1973F
SMAD2 p.S464X
BVR-O-04
BRAF p.N581S
APC p.E941X
APC p.Q1447X
FANCC p.S119X
KRAS p.L19F
PIK3R1 p.R461X
PIK3R1 p.M326_G336delinsISLQ
PTEN p.R233X
TP53 p.V157_A159del
BVR-O-05
BRAF p.D594N
ARID1A p.E1776X
PBRM1 p.R1160X
PIK3CA p.E542K
PIK3CA p.R93Q
RNF43 p.W15X
RNF43 p.E258X
TGFBR2 p.R528G
BVR-O-06
BRAF p.D594G
APC p.R876X
APC p.E1552X
EP300 p.D452X
NRAS p.G13D
TP53 p.R342X
CCNE1 AMP
BVR-O-15
BRAF p.D594H
APC p.S1415X
APC p.R213X
MLH1 c.844+1G>A
RAF1 p.S257L
TET2 pQ604X
TP53 p.Q204X
WT1 p.P41X
MGA p.S879X
RANBP2 p.G2325X

Figure S9

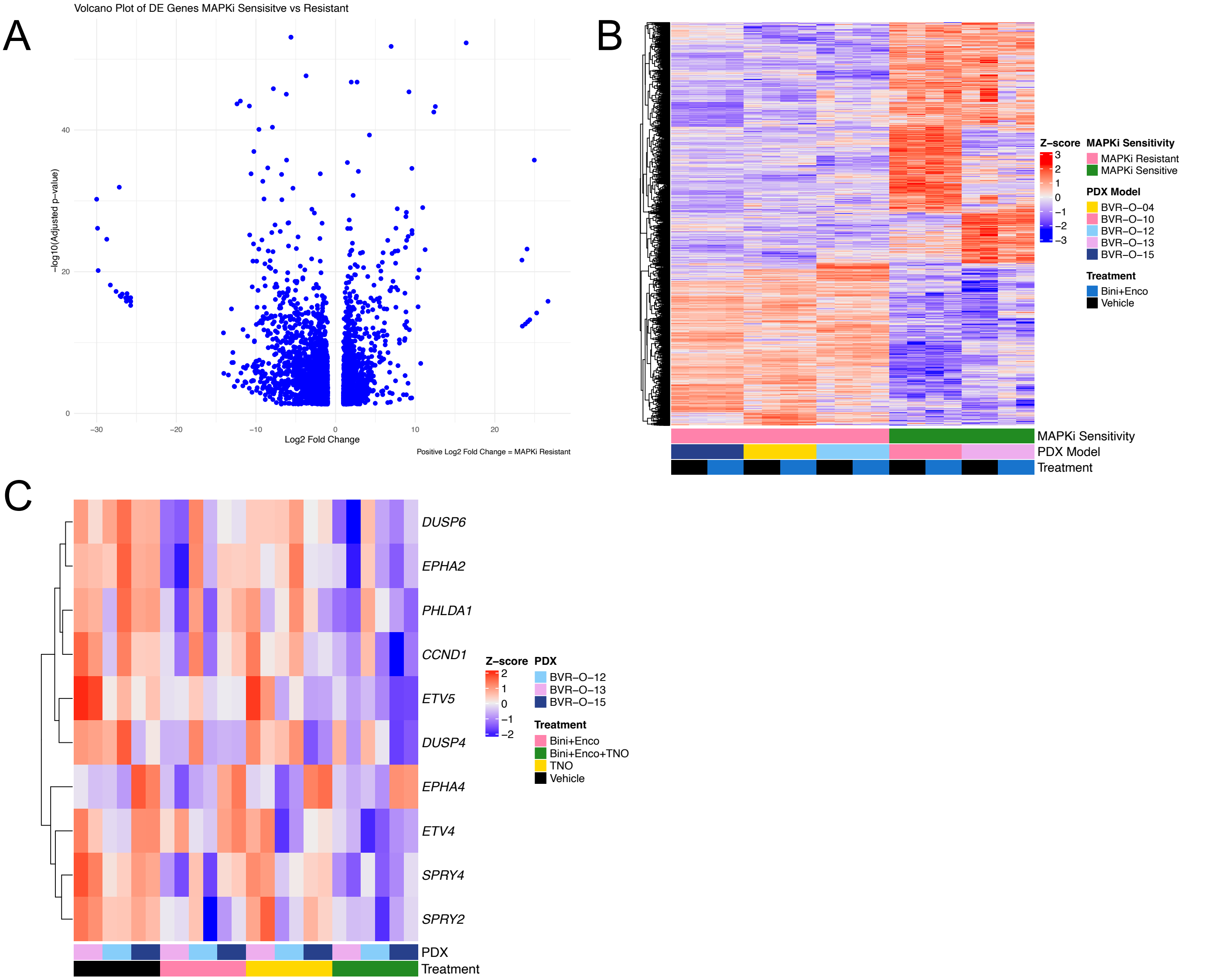
A
Volcano Plot of DE Genes MAPKi Sensisitve vs Resistant
−log10(Adjusted p−value)
0
20
40
−30
−20
−10
0
10
20
Log2 Fold Change
Positive Log2 Fold Change = MAPKi Resistant
B
Z−score
3
2
1
0
−1
−2
−3
MAPKi Sensitivity
MAPKi Resistant
MAPKi Sensitive
PDX Model
BVR−O−04
BVR−O−10
BVR−O−12
BVR−O−13
BVR−O−15
Treatment
Bini+Enco
Vehicle
MAPKi Sensitivity
PDX Model
Treatment
C
DUSP6
EPHA2
PHLDA1
CCND1
ETV5
DUSP4
EPHA4
ETV4
SPRY4
SPRY2
Z−score
2
1
0
−1
−2
PDX
BVR−O−12
BVR−O−13
BVR−O−15
Treatment
Bini+Enco
Bini+Enco+TNO
TNO
Vehicle
PDX
Treatment

**Figure S10**

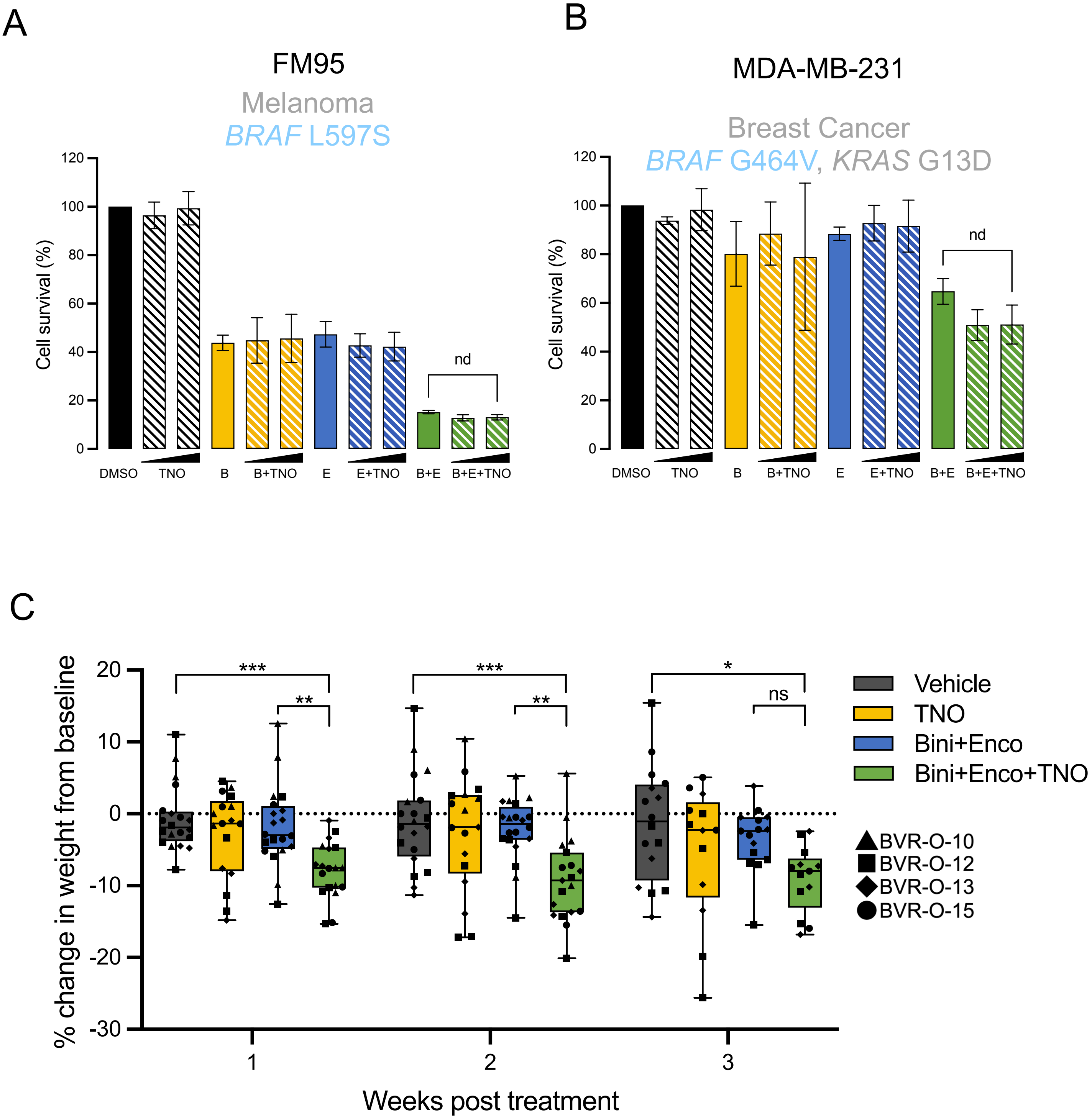

A
FM95
Melanoma
BRAF L597S
Cell survival (%)
nd
DMSO
TNO
B
B+TNO
E
E+TNO
B+E
B+E+TNO
B
MDA-MB-231
Breast Cancer
BRAF G464V, KRAS G13D
Cell survival (%)
nd
DMSO
TNO
B
B+TNO
E
E+TNO
B+E
B+E+TNO
C
% change in weight from baseline
Weeks post treatment
***
**
***
**
*
ns
Vehicle
TNO
Bini+Enco
Bini+Enco+TNO
BVR-O-10
BVR-O-12
BVR-O-13
BVR-O-15


**Figure S11**
