## Supplemental text for "CDK4/6 and SHP2 mediate BRAF/MEK inhibitor resistance in Class 2 and 3 BRAF mutant cancers"

**Supplemental Methods:**

**Antibodies used & dilutions**

| **Protein** | **Company & product number** | **Species** | **Dilution** |
| --- | --- | --- | --- |
| p44/42 MAPK (Erk1/2) (137F5) | Cell Signaling #4695S | Rabbit | 1:1000 |
| pERK (Phospho-p44/42 MAPK (Erk1/2) (Thr202/Tyr204) (D13.14.4E) XP®) | Cell Signaling #4370s | Rabbit | 1:1000 |
| pRb 807 | Cell Signaling #9308 | Rabbit | 1:1000 |
| Rb (Recombinant Anti-Rb antibody [EPR17512]) | Abcam #ab181616 | Rabbit | 1:4000 |
| Rb | Cell Signaling #9309 | Mouse | 1:1000 |
| CDK2 (E8J91) XP(R) | Cell Signaling #18048 | Rabbit | 1:1000 |
| CDK4 (D9G3E) | Cell Signaling #12790 | Rabbit | 1:1000 |
| CDK6 | Abcam #AB124821 | Rabbit | 1:1000 |
| CDK6 | Santa Cruz #SC-7961 | Mouse | 1:1000 |
| Cyclin D1 (E3P5S) XP(R) | Cell Signaling #55506 | Rabbit | 1:1000 |
| Cyclin E1 (HE12) | Cell Signaling #4129 | Mouse | 1:1000 |
| Beta-actin | Cell Signaling #4970 | Rabbit | 1:5000 |
| Vinculin (E1E9V) | Cell Signaling #13901 | Rabbit | 1:1000 |
| Alpha-Tubulin | Sigma Aldrich #T5168 | Mouse | 1:5000 |
| SHP2 | Cell Signaling #3397 | Rabbit | 1:1000 |
| STAT1 (D1K9Y) | Cell Signaling #14994S | Rabbit | 1:1000 |
| Phospho-Stat1 (Tyr701) | Cell Signaling #9167S | Rabbit | 1:1000 |
| Anti-Mouse IgG HRP-linked Antibody | Cell Signaling #7076 | Horse | 1:5000 |
| Anti-Rabbit IgG-HRP Conjugate | BioRad #1706515 | Goat | 1:5000 |

**Immunoblotting of functional genomics experiments**

MDA-MB-231 (transfected with non-targeting, or *CDK4*-targeting, or *CDK6*-targeting siRNA) and NCI-H1666 (transduced with non-targeting gRNA or *PTPN11*-targeting gRNAs) cell pellets were harvested, washed in PBS, and stored in -80 °C until processed for protein extraction. Cells were lysed on ice for 15 min using ice-cold radioimmunoprecipitation assay buffer (1% Nonidet P-40, 150 mM NaCl, 5 mM EDTA, 50 mM Tris (pH 7.5), 0.5% deoxycholic acid, 0.1% SDS) supplied with 1× protease inhibitor cocktail (Roche, cat. no. 11836153001), NaF (5 mM), and Na3VO4 (1 mM). Standard SDS-PAGE protein separation protocol was performed, and proteins were transferred to methanol-activated Polyvinylidene fluoride (PVDF) membranes using the TransBlot Turbo transfer system, as per the manufacturer’s protocol (BioRad). Membranes were blocked for one hour in Tris-Buffered Saline (TBS) solution containing 0.1% Tween-20 (TBS-T) and 1% bovine serum albumin (Bio Basic). For FLAG-Cas9 blots, after blocking, membranes were incubated with monoclonal anti-FLAG M2-horseradish peroxidase (HRP) (1:8000 dilution; Sigma, cat. no. A8592) in 1% BSA-supplemented TBS-T for one hour. Membranes were then washed with TBS-T three times, 5 minutes each, before incubation with enhanced chemiluminescence reagent (ZmTech Scientifique) for 1 minute.

**WES and RNAseq Analysis**

WES was performed on genomic DNA from all patient-derived xenografts and cell lines used through Novogene’s (Sacramento, CA, USA) WES pipeline as follows. Genomic libraries were prepared using 400 ng of genomic DNA with a SureSelect Human All Exon V6 capture kit (Agilent, Santa Clara, CA, USA) and sequenced with the NovaSeq X Plus or NovaSeq 6000 platform (Illumina, San Diego, CA, USA). The genome dataset was aligned with the human genome GCRh38 and annotated with ANNOVAR. RNA counts were obtained through Novogene’s RNAseq pipeline as follows. Messenger RNA was purified from total RNA using poly-T oligo-attached magnetic beads. cDNA was synthesized using random hexamer primers and either dUTP or dTTP based on the library. Quantified libraries were pooled and sequenced on Illumina platforms and paired-end reads were generated. Hisat2 v2.0.5 were used to build the reference genome and align the paired-end reads. Gene expression was obtained with featureCounts v1.5.0-p3. The MAPK Pathway Activation Score (MPAS) score was calculated as previously described [1] by adding the Z-scores for each of the 10 genes of the MPAS gene signature and dividing the total by 10, the number of genes included in the gene signature. The same methodology was applied to create the E2F Targets and G2M Checkpoint scores, each derived from the corresponding Hallmark MSigDB gene sets [2] and by dividing the number of genes in the respective gene sets. Statistical analysis was performed with One-way ANOVA and Tukey’s multiple comparisons test.

**Cell-cycle analysis**

Cell cycle analysis was performed by plating 350,000-500,000 cells per well of a 6-well plate for a total of ~1,000,000 cells per condition. 4-6hrs later, cells were synchronized by aspirating media, washing 2X with PBS and incubated in low serum conditions (0.5% FBS) overnight. The following day, cells were treated with inhibitors for 24hrs and then washed twice with ice-cold PBS containing 1% FBS. Cells were stained with 50 mg/mL propidium iodide solution in hypotonic buffer (0.1% Triton X-100 and 0.1% sodium citrate) for at least 20 minutes in the dark. A minimum of 20,000 cycling cells were analyzed using BD FACSCanto II flow cytometer (REF 338960). Data was analyzed using ModFit LT for Windows software (v 4.1.7). Diploid cell cycle phases were used for analyses.

**Apoptosis Assays**

Drug-induced apoptosis was assessed by annexin V combined with propidium iodide (PI) staining. Cells and supernatants were collected following 72hr treatment and stained with Annexin-V-FITC Detection kit (BD Pharmingen, cat. no. 560931) and measured on a BD FACSCanto II flow cytometer.

**Proliferation assays**

*siRNA transfection experiment*

The IncuCyte software was used to define the percentage of cell confluence in each condition and the relative change in confluence at 72hrs (endpoint) in reference to the earliest time point was calculated. The resulting values for all the experimental conditions were normalized to the cells transfected with non-targeting siRNA and treated with DMSO (the reference condition). Values from 3 independent biological replicates were combined and plotted in GraphPad Prism. Two-way ANOVA test (multiple comparisons; Tukey correction) was used to compare different conditions and the resulting p-values are stated on the graph.

*Two-color competition assay*

The IncuCyte Software was used to quantify the green (GFP-positive) and orange (mCherry-positive) cells in every experimental condition. A stringent analysis threshold was used to eliminate objects that are positive for the two colors, hence ensuring accurate quantifications. The percentage of GFP-positive cells from the total population was then calculated for the D0 and D7 samples. The relative change in this percentage was calculated for different experimental conditions (NT gRNA, *PTPN11* gRNA1, *PTPN11* gRNA2), and plotted using GraphPad Prism (GraphPad Software, San Diego CA). Different conditions were compared using a two-tailed t-test after combining the results of 3 biological replicates (from three independent gRNA transductions) and the resulting *p*-values are stated on the graph.

gRNAs sequences

| Gene/region | Sequence |
| --- | --- |
| *PTPN11* | gRNA1: GAGACTTCACACTTTCCGTT  gRNA2: TACAGTACTACAACTCAAGC |
| Non-targeting | CTGAAA AAGGAAGGAGTTGA |
| *AAVS1* | ATCCTGTCCCTAGTGGCCC |

**Supplemental Tables**

**Table S1: Characteristics of patients enrolled on the BEAVER trial**

| **Characteristic** | **Total n (%)** |
| --- | --- |
| **Median age (years, range)** | 59 (40-73) |
| **Gender** |  |
| Female | 15 (65%) |
| Male | 8 (35%) |
| **Median number of prior systemic therapies**  **(range)** | 1 (0-6) |
| 0 | 2 |
| 1 | 11 |
| 2+ | 10 |
| **Cancer Type** |  |
| Melanoma | 6 (26%) |
| Colorectal | 6 (26%) |
| Pancreaticobiliary - cholangiocarcinoma | 3 (13%) |
| Pancreaticobiliary - ampullary | 1 (4%) |
| Pancreaticobiliary – gall bladder | 1 (4%) |
| Pancreaticobiliary – pancreatic | 1 (4%) |
| NSCLC | 2 (9%) |
| Breast | 1 (4%) |
| Uterine | 1 (4%) |
| Small Bowel | 1 (4%) |
| **BRAF mutation class** |  |
| Class 1 | 1 (4%) |
| Class 2 | 9 (39%) |
| Class 3 | 13 (57%) |

**Table S2: Treatment related adverse events**

| **Adverse Event** | **Grade 1-2** | **Grade 3** |
| --- | --- | --- |
| ANY | 19 (83%) | 5 (22%) |
| BLURRED VISION | 13 (57%) | 0 |
| NAUSEA | 11 (48%) | 1 (4%) |
| VOMITING | 11 (48%) | 0 |
| FATIGUE | 10 (43%) | 1 (4%) |
| DIARRHEA | 6 (26%) | 0 |
| CREATININE INCREASED | 5 (22%) | 0 |
| ALKALINE PHOSPHATASE INCREASED | 4 (17%) | 0 |
| CPK INCREASED | 4 (17%) | 0 |
| ALANINE AMINOTRANSFERASE INCREASED | 3 (13%) | 0 |
| ANOREXIA | 3 (13%) | 0 |
| CHILLS | 3 (13%) | 0 |
| CONSTIPATION | 3 (13%) | 0 |
| DYSPNEA | 3 (13%) | 0 |
| FEVER | 3 (13%) | 0 |
| ABDOMINAL PAIN | 2 (9%) | 0 |
| ASPARTATE AMINOTRANSFERASE INCREASED | 2 (9%) | 1 (4%) |
| EDEMA LIMBS | 2 (9%) | 0 |
| PRURITUS | 2 (9%) | 0 |
| RETINOPATHY | 2 (9%) | 0 |
| SERUM AMYLASE INCREASED | 2 (9%) | 0 |
| ABDOMINAL DISTENSION | 1 (4%) | 0 |
| LIPASE INCREASED | 1 (4%) | 1 (4%) |
| RASH MACULO-PAPULAR | 1 (4%) | 1 (4%) |
| GASTROINTESTINAL DISORDERS - OTHER | 1 (4%) | 0 |
| ALOPECIA | 1 (4%) | 0 |
| CONFUSION | 0 (0%) | 1 (4%) |

**Table S3: Characteristics of patients and biopsies used for PDX development**

| Patient ID | Cancer Type | Patient Gender | *BRAF* mutation | *BRAF* mutation class | Pre-treatment biopsy | On-treatment biopsy | | PDX attempted/established | Bini+Enco sensitive or resistant *in vivo* |
| --- | --- | --- | --- | --- | --- | --- | --- | --- | --- |
| BVR-M-02 | Melanoma | Male | V600G | Class 1 | Yes | No | Yes/Yes | | resistant |
| BVR-M-03 | Melanoma | Male | G469S | Class 2 | No | No | N/A | | N/A |
| BVR-M-04 | Melanoma | Female | G466E* | Class 3 | Yes | Yes | Yes/Yes | | resistant |
| BVR-M-05 | Melanoma | Female | K601E | Class 2 | No | No | N/A | | N/A |
| BVR-M-07 | Mucosal Melanoma | Female | G469A | Class 2 | No | No | N/A | | N/A |
| BVR-M-08 | Melanoma | Male | D594G | Class 3 | No | No | N/A | | N/A |
| BVR-O-01 | Breast | Female | K601E | Class 2 | Yes | No | Yes/No | | N/A |
| BVR-O-02 | NSCLC | Male | N581I* | Class 3 | Yes | No | Yes/No | | N/A |
| BVR-O-03 | Uterine | Female | SLC45A3 exon1_ BRAF exon8 Fusion | Class 2 | No | No | N/A | | N/A |
| BVR-O-04 | Colorectal | Female | N581S | Class 3 | Yes | No | Yes/Yes | | resistant |
| BVR-O-05 | Gallbladder | Female | D594N | Class 3 | No | No | N/A | | N/A |
| BVR-O-06 | Colorectal | Female | D594G | Class 3 | No | No | N/A | | N/A |
| BVR-O-07 | Ampullary | Female | D594G | Class 3 | No | No | N/A | | N/A |
| BVR-O-09 | Pancreatic | Female | M484_P490delinsIAR | Class 2 | Yes | Yes | Yes/No | | N/A |
| BVR-O-10 | Cholangiocarcinoma | Female | N581I | Class 3 | Yes | Yes | Yes/Yes | | sensitive |
| BVR-O-11 | NSCLC | Female | N581I* | Class 3 | Yes | No | Yes/No | | N/A |
| BVR-O-12 | Colorectal | Male | G466E | Class 3 | Yes | Yes | Yes/Yes | | resistant |
| BVR-O-13 | Colorectal | Female | D594N | Class 3 | Yes | Yes | Yes/yes | | sensitive |
| BVR-O-14 | Small Bowel | Male | D594N | Class 3 | No | No | N/A | | N/A |
| BVR-O-15 | Colorectal | Female | D594H | Class 3 | Yes | Yes | Yes/Yes | | resistant |
| BVR-O-16 | Cholangiocarcinoma | Female | G469A | Class 2 | No | No | N/A | | N/A |
| BVR-O-17 | Colorectal | Male | L597R | Class 2 | Yes | Yes | Yes/Yes | | resistant |
| BVR-O-18 | Cholangiocarcinoma | Male | K601E | Class 2 | Yes | Yes | Yes/Yes | | resistant |

*BRAF mutation was identified and reported in clinical sequencing but was not identified in subsequent tissue analyzed using the OCAv3 or TSO500 targeted sequencing panels.

**Supplemental Figure Legends**

**Supplemental Figure 1: PFS and OS of BEAVER cohort**

**A)** Median PFS for the entire cohort was 2.3 months **B)** Median OS for the entire cohort was 6.2 months.

**Supplemental Figure 2: BEAVER trial response data by BRAF Class**

**A)** No significant differences in patient response by BRAF Class. Fisher’s exact test, P=0.7684 **B)** No significant difference in PFS by BRAF Class. Log rank test, P=0.521 **C)** No significant difference in OS by BRAF Class. Log rank test, P=0.786

**Supplemental Figure 3: BEAVER trial response data by tumour type**

**A)** Melanoma and pancreaticobiliary patients had better responses to Bini+Enco targeted therapy when compared to patients with colorectal and other cancer types. Fisher’s exact test, P=0.0081 **B)** BEAVER trial patients with non-V600E BRAF mutant colorectal and other cancer types had worse PFS than melanoma and pancreaticobiliary. Log rank test, P=0.02 **C)** No significant difference in OS by cancer type. Log rank test, P=0.359 **D)** Patient response stratified by cancer type and *TP53* mutation status.

**Supplemental Figure 4: BEAVER PDX response and profile**

**A)** Patients whose tumors had larger change in target lesion size were significantly more likely to result in an engraftment of the PDX (P=0.0336; Mann-Whitney test). **B)** Genomic profile of patients (blue) and corresponding PDXs (red). **C)** Tumour growth curves of PDX response to Bini+Enco treatment over 14-21 days.

**Supplemental Figure 5: Characterization of BRAF Class 2 Bini+Enco resistant cells**

**A)** IC50 to combined Bini+Enco treatment for all 4 parental/resistant pairs. No IC50 value could be achieved for the 22RV1 resistant cells. **B)** Cells were treated for 24hrs with 200nM Binimetinib + 1000nM Encorafenib and apoptosis levels were measured. Apoptosis assay revealed Class 2 BRAF resistant cells have a protective effect against Bini+Enco induced apoptosis vs. parental cells. Bini+Enco treatment induced apoptosis in all 4 parental cells. Brown-Forsythe and Welch ANOVA (Dunnett’s T3 multiple comparisons test) *P<0.05, **P<0.01, ***P<0.001, ****P<0.0001.

**Supplemental Figure 6: Evaluating cell cycle alterations in non-V600E BRAF mutant tumors**

**A)** MPAS, E2F, G2M RNAseq Scores for MDA-MB-231, HMV-II, FM95 and 22RV1 parental/resistant cells treated with DMSO or Bini+Enco. One-way ANOVA. **B)** Gene expression of the 10 MPAS genes by cell line **C)** MPAS, E2F and G2M RNAseq scores for parental melanoma (FM95 & HMV-II) and non-melanoma (MDA-MB-231, 22RV1) cell lines treated with DMSO or Bini+Enco for 24hrs. One-way ANOVA *P<0.05, **P<0.01, ***P<0.001, ****P<0.0001. **D)** Representative flow cytometry plots for the cell cycle analysis for each cell line and condition.

**Supplemental Figure 7: Evaluating the efficacy and safety of Bini+Enco+Palbo triple therapy in non-V600E BRAF mutant models**

**A)** Clonogenic assay in the Class 3 BRAF mutant H508 cell line with DMSO, Enco, Bini, and Palbo treatment. Triple therapy significantly more effectively reduces cancer cell growth vs. Bini+Enco (Unpaired t-test B+E+P vs. B+E). **B)** Tumor growth curve for Class 3 BRAF BVR-O-04, BVR-O-10, BVR-O-12, and BVR-O-13 PDX models. PDXs treated with vehicle, binimetinib (15mg/kg/d) + encorafenib (75mg/kg/d) (bini + enco) or palbociclib (80mg/kg/day) (palbo). The drug doses for the triple therapy groups were: binimetinib (15mg/kg/d) + encorafenib (75mg/kg/d) + palbociclib (80mg/kg/day) (B+E+P) for PDXs BVR-O-04/10/12 and binimetinib (15mg/kg/d) + encorafenib (50mg/kg/d) + palbociclib (80mg/kg/day) (B+E+P) for PDXs BVR-O-13. Bini+Enco+Palbo triple therapy significantly inhibits tumour growth *in vivo* in all models except BVR-O-12. One-way ANOVA on AUC *P<0.05, **P<0.01, ***P<0.001, ****P<0.0001. **C)** Immunoblot of tumours from BVR-O-04 taken at endpoint. Each lane represents protein lysate from a different biological replicate (tumor). **D)** Mice weight over treatment period by treatment group and PDX model. No significant changes in weight were observed in vehicle vs B+E+P or B+E vs B+E+P treated mice. Three B+E+P mice (GCRC-Mel1) died on Day 19; these deaths are suspected to have occurred due to technical issues with gavage. For BVR-O-17, one vehicle-treated mouse reached a humane end-point for tumor size on Day 14. For BVR-O-04, two vehicle-treated mice, one palbo treated mouse, and one bini+enco treated mouse reached the human end-point for tumor size on day 24.

**Supplemental Figure 8: Evaluating MAPK pathway and cell cycle transcriptomics in B+E+P PDXs**

**A)** MPAS heatmap for GCRC-Mel1, BVR-O-04, and BVR-O-17 treated with Vehicle, Bini+Enco, Palbo, and Bini+Enco+Palbo. **B)** E2F hallmark gene set heatmap for GCRC-Mel1, BVR-O-04, and BVR-O-17 treated with Vehicle, Bini+Enco, Palbo, and Bini+Enco+Palbo. **C)** G2M hallmark gene set heatmap **f**or GCRC-Mel1, BVR-O-04, and BVR-O-17 treated with Vehicle, Bini+Enco, Palbo, and Bini+Enco+Palbo.

**Supplemental Figure 9: Analysis of circulating tumor DNA (ctDNA) from patients enrolled on the BEAVER trial**

All patients enrolled in the BEAVER trial provided serial blood samples prior to starting treatment and periodically during treatment. Plasma was collected and ctDNA was analyzed using the TSO500 ctDNA v2 assay. For patients who experienced tumor regression, we analyzed samples that were collected prior to treatment, prior to cycle 2, day 1, and at the time of progression. For patients who did not experience tumor regression, we analyzed ctDNA samples that were collected prior to treatment and at the time of progression. **A)** Results from patients with Class 2 *BRAF* mutations **B)** Results from patients with Class 3 *BRAF* mutations. New mutations that were only present in samples taken at the time of PD are highlighted with a box. VAF = variant allele frequency; C1D1 = cycle 1, day 1.

**Supplemental Figure 10: Transcriptomic profiling of Class 3 BRAF mutant PDXs**

**A)** Volcano plot of differentially expressed genes (baseMean > 50, absolute Log2FoldChange >1, padj < 0.05) between MAPKi sensitive and resistant PDX. 3651 genes total, 1469 genes overexpressed in MAPKi resistant PDXs (Vehicle & Bini+Enco), 2182 genes overexpressed in MAPKi sensitive PDXs (Vehicle & Bini+Enco). **B)** Heatmap of the 3651 differentially expressed genes in MAPKi sensitive (BVR-O-10 & BVR-O-13) and resistant (BVR-O-04, BVR-O-12 & BVR-O-15) PDX. **C)** GSEA analysis of Bini+Enco and Vehicle treated PDX grouped by MAPKi sensitivity, plot shows enriched gene sets in MAPKi Resistant PDXs. **C)** Heatmap of the 10 MPAS genes from BVR-O-12, BVR-O-13 & BVR-O-15 PDX models that were treated with vehicle, bini+enco, TNO155 or B+E+T.

**Supplemental Figure 11: Evaluation of BRAF/MEK/SHP2 inhibition *in vitro* and *in vivo***

Quantification of clonogenic assays of cancer cells with Class 2 *BRAF* mutations that were treated with combinations of BRAF, MEK and SHP2 inhibitors, encorafenib, binimetinib and TNO155 respectively in **A)** FM95 (melanoma) and **B)** MDA-MB-231 (breast cancer) cells. Drug doses: FM95 - Bini 20nM, Enco 100nM, TNO 100nM and 1µM; MDA-MB-231 - Bini 40nM, Enco 200nM, TNO 100nM and 1µM. N=3 biological replicates plotted. One-way ANOVA *P<0.05, **P<0.01, ***P<0.001, ****P<0.0001. **C)** Mice weight over treatment period by treatment group and PDX model. The B+E+T triple therapy was associated with more weight loss in mice compared to vehicle or B+E alone. Two-way ANOVA *P<0.05, **P<0.01, ***P<0.001, ****P<0.0001. Mice from BVR-O-10, BVR-O-12 and BVR-O-13 PDXs were treated with Binimetinib (15mg/kg/d) + Encorafenib (75mg/kg/d), while those from BVR-O-15 was treated with Binimetinib (15mg/kg/d) + Encorafenib (50mg/kg/d).
